# Functional filter for whole genome sequence data identifies stress impact, non-coding alternate polyadenylation site variants >5kb from coding DNA

**DOI:** 10.1101/2023.05.10.23289736

**Authors:** Sihao Xiao, Zhentian Kai, Daniel Murphy, Dongyang Li, Dilip Patel, Adrianna Bielowka, Maria E. Bernabeu-Herrero, Awatif Abdulmogith, Andrew D Mumford, Sarah Westbury, Micheala A Aldred, Neil Vargesson, Mark J Caulfield, Genomics England Research Consortium, Claire L Shovlin

## Abstract

Despite whole genome sequencing (WGS), why do many single gene disorder cases remain unsolved, impeding diagnosis and preventative care for people whose disease-causing variants escape detection? Early WGS data analytic steps prioritize protein-coding sequences. To simultaneously prioritise variants in non-coding regions rich in transcribed and critical regulatory sequences, we developed GROFFFY, an analytic tool which integrates coordinates for regions with experimental evidence of functionality. Applied to WGS data from solved and unsolved hereditary hemorrhagic telangiectasia (HHT) recruits to the 100,000 Genomes Project, GROFFFY-based filtration reduced the mean number of variants per DNA from 4,867,167 to 21,486, without deleting disease-causal variants. In three unsolved cases (two related), GROFFFY identified ultra-rare deletions within the 3’ untranslated region (UTR) of the proto-oncogene *SMAD4*, where germline loss-of-function alleles cause combined HHT and colonic polyposis. Sited >5.4kb distal to coding DNA, the deletions did not modify or generate microRNA binding sites, but instead disrupted the sequence context of the final cleavage and polyadenylation site necessary for protein production: By iFoldRNA, an AAUAAA-adjacent 16 nucleotide deletion brought the cleavage site into inaccessible neighboring secondary structures, while a 4-nucleotide deletion unfolded the downstream RNA polymerase II roadblock. Monocyte *SMAD4* RNA expression differed between patients and controls in resting and cycloheximide-stressed states. Patterns predicted the mutational site for an unrelated case, where a complex insertion was subsequently identified. In conclusion, a new type of functional rare variant is described, exposing novel regulatory systems based on polyadenylation. Extension of coding sequence-focused gene panels is required to capture these variants.

## INTRODUCTION

Whole genome sequencing (WGS) is an established component of medical genetic and research repertoires, but currently, the majority of its potential is unrealized. In any one individual, WGS identifies millions of DNA variants compared to reference sequences. These are present in ∼20,000 protein-coding genes, and also in much less understood regions of the genome that have diverse functions including transcription into noncoding RNAs, participation in DNA chemical changes that modify transcription, and binding to other nucleic acids or proteins (*1,2*).

Current WGS clinical foci are almost exclusively on a subgroup of protein-coding genes where biological function is already known. In research spheres, in order to reduce the number of variables per sample, interrogation of WGS data also commences with prioritization methods, usually based on selection of specific genomic regions. Variants in the non-coding genome, while not pre-depleted by the sequencing methodology, are effectively deleted in the early analytic stages of variant prioritization. Importantly, application of these WGS methods leave large proportions of patients unsolved, without a genetic diagnosis (*3*).

There is no accurate map of all functional genomic regions in human genomes, and it is difficult to predict *a priori,* where all regulatory elements for a specific gene locus would be located. We hypothesized however, that it would be possible to design a more efficient variant prioritization method for WGS because markers of epigenetics and DNA-protein interactions have been applied genome-wide by molecular laboratories, and an enormous body of biological experimental data made publicly available. As a result, there now exist repositories of information indicating which sections of DNA are more or less likely to have a functional role in at least one examined tissue.

We designed a <u>g</u>enomic <u>r</u>egions <u>o</u>f functionality filter for priorit<u>y</u> (*GROFFFY)* based on published experimental data particularly from ENCODE (*4–6*) and performed validation and discovery analyses in WGS data from patients recruited to the 100,000 Genomes Project (*7*). To accelerate clinical impact, we focused discovery analyses on noncoding regions of a proto-oncogene examined in diagnostic and screening gene panels. *SMAD4* is ubiquitously expressed and encodes the common partner SMAD which regulates signaling by transforming growth factor (TGF)-β, bone morphogenetic protein (BMP), and activin ligands (*8,9*). As indicated by its function and earlier gene names (*DPC4*-deleted in pancreatic cancer 4; *MADH4-*mothers against decapentaplegic), the SMAD4 protein has major pathological and developmental roles (*8,9*). *SMAD4* is a target of cancer genetic diagnostics because it is a driver gene for major cancers due to somatic loss (*8,9*), and because germline heterozygous loss causes gastrointestinal polyposis (“juvenile polyposis”/JP) where untreated hamartomatous polyps can undergo malignant transformation leading to colon, gastric and other cancers (*8,9*). Heterozygous loss also causes TGF-β/BMP-related vasculopathies including hereditary hemorrhagic telangiectasia (HHT) which usually results from a loss-of-function variant in *ACVRL1* or *ENG* (*10*), but where identification of combined HHT/JP due to *SMAD4* (*11,12*) allows patients to benefit from life-long polyposis and aortopathy screening programs (*9*). Scientifically, *SMAD4* is of great interest because despite its wide-ranging roles in development and disease, little is known of its regulation (*8,9*).

Here we report that this new WGS analytic approach identifies a novel type of functional DNA variant uncaptured by usual clinical sequencing methodologies.

## RESULTS

### GROFFFY defines biologically validated regions of functionality

By only including regions where biological experiments have generated evidence in favor of functional roles, GROFFFY essentially excludes biologically less important regions of the genome. Nevertheless, the GROFFFY filter region based on positive selection of transcribed loci and candidate regulatory element (cREs), and masking of repetitive regions, included 44.4% of the human genome. A heatmap at 500kb resolution is provided in Fig. 1A. A more detailed view of GROFFFY is provided in Fig. 1B.

**Fig. 1:**
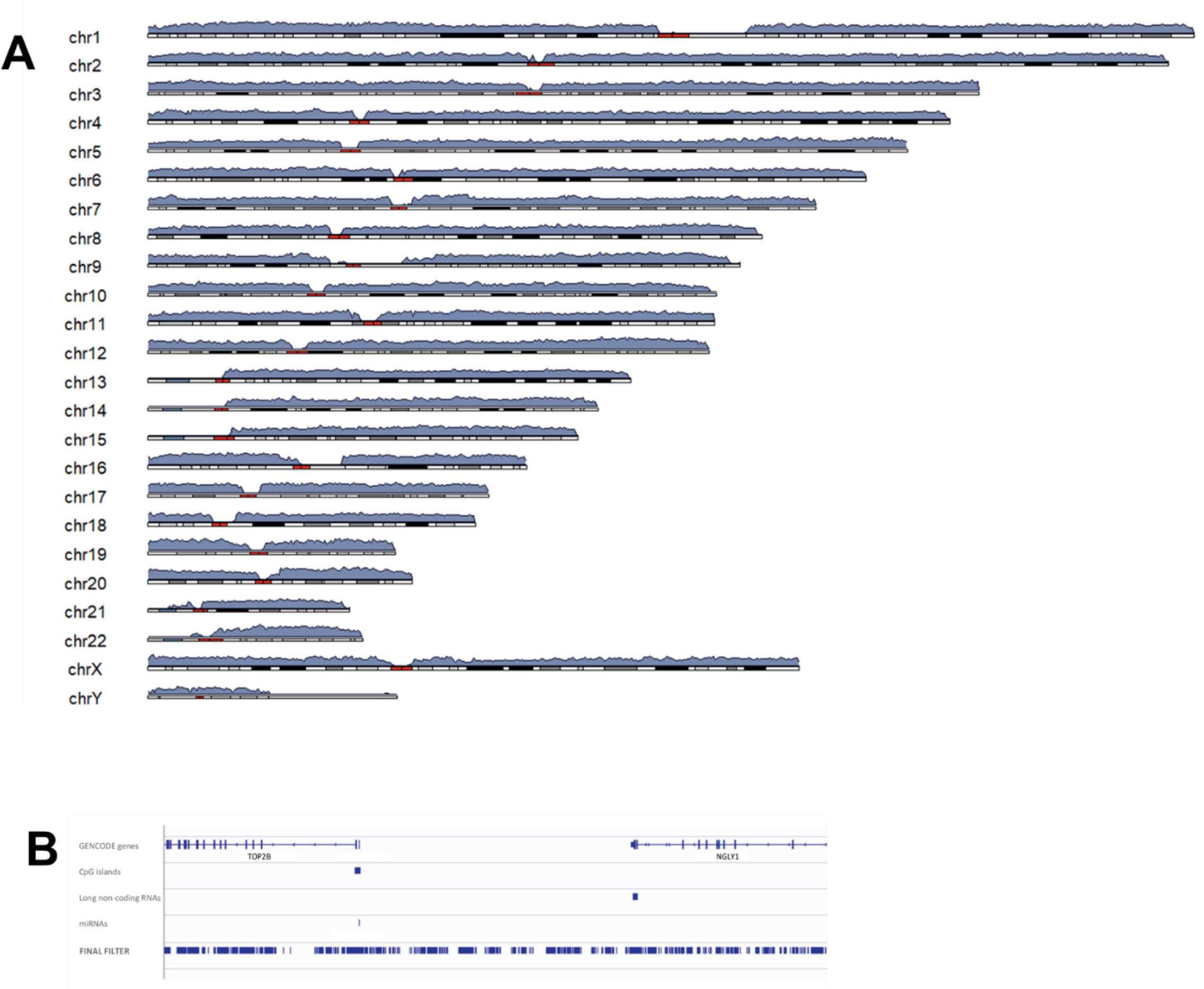
GROFFFY and the human genome. **A)** Heatmap displaying GROFFFY categorization of Genome Reference Consortium Human [GRCh] Build 38 (*13*). The heatmap maps 61,799 data points at 500kb resolution, and heights represent the percentage of each 500kb region included in GROFFFY. **B)** Higher resolution image from a randomly chosen region of the genome (on chromosome 3: chr3:25,597,986-2-25,783,443). The top 4 tracks illustrate sources, from top: GENCODE (*6*) gene annotations, CpG islands (*14,15*), long non-coding RNAs (*16,17*) and miRNAs (*18*). The lowest track illustrates the final filter. Note that this filter contains both intra and intergenic regions for the region, and that the raw data were not subjected to any processed annotation tracks.

### GROFFFY substantially reduces the number of DNA variants per DNA

The scale of the bioinformatics challenge was emphasized by the pre-filtration number of DNA variants per individual which ranged from 4,786,039 to 5,070,340 (mean 4,867,167). Applying GROFFFY as a first filter reduced the mean number of variants by 2,812,015 (Fig. 2A, Fig. 2B). Restricting to rare variants with population allele frequencies <2x10^-4^ (*19*) removed means of 2,476,589 (*20*) and 2,483,377 (*21*) variants/DNA according to database (*20,21*) (Fig. 2A, Fig. 2B). After removing variants with a Combined Annotation-Dependent Depletion (CADD) (*22*) score <10, the mean number of unique, rare and impactful DNA variants per genome was 21,486 (Fig. 2C).

**Fig. 2:**
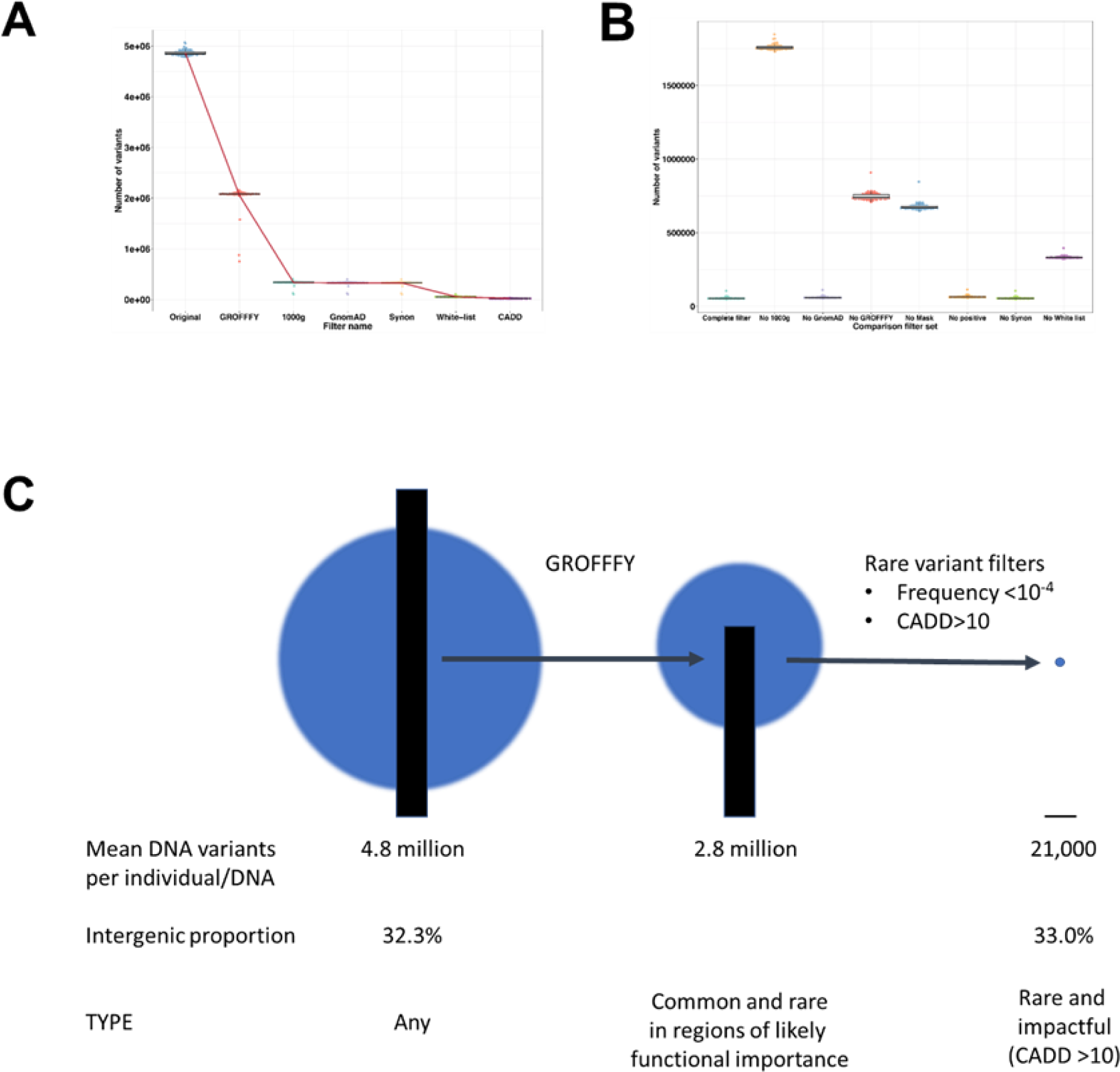
Application of GROFFFY to whole genome sequences. **A)** Serial application of GROFFFY; allele frequency filters based on frequencies in the 1000 Genomes (1000g) (*20*) or GnomAD (*21*); synonymous (Synon.) filter, and white-listed filter (see Methods and *Tables S1-S4* for further details). Where error bars are not visible at the illustrated scale, exact numeric data are provided in *Table S5***. B)** Number of variants remaining per DNA after applying each comparator filter set. **C)** Number, site and type of DNA variants present in 98 human whole genomes before and after application of GROFFFY and other filters, scaled in one dimension (black bars) and two dimensions (blue circles). CADD, combined annotation dependent depletion score where >10 represents a variant in the top 10% of deleteriousness (*22*). Irrespective of other filters applied, GROFFFY, and its individual components, significantly reduced the number of variants compared to the other tested filter sets (*Fig. S1, Fig. S2*).

GROFFFY did not delete key variants, as shown by the validation dataset: All already-known pathogenic variants in the unfiltered dataset were retained post filtration (Fig. 3A). Further, in the discovery set of 98 whole genomes, for *ACVRL1* and *ENG,* the majority of identified novel variants clustered to the exons and flanking regions sequenced in clinical diagnostics (Fig. 3B).

**Fig. 3:**
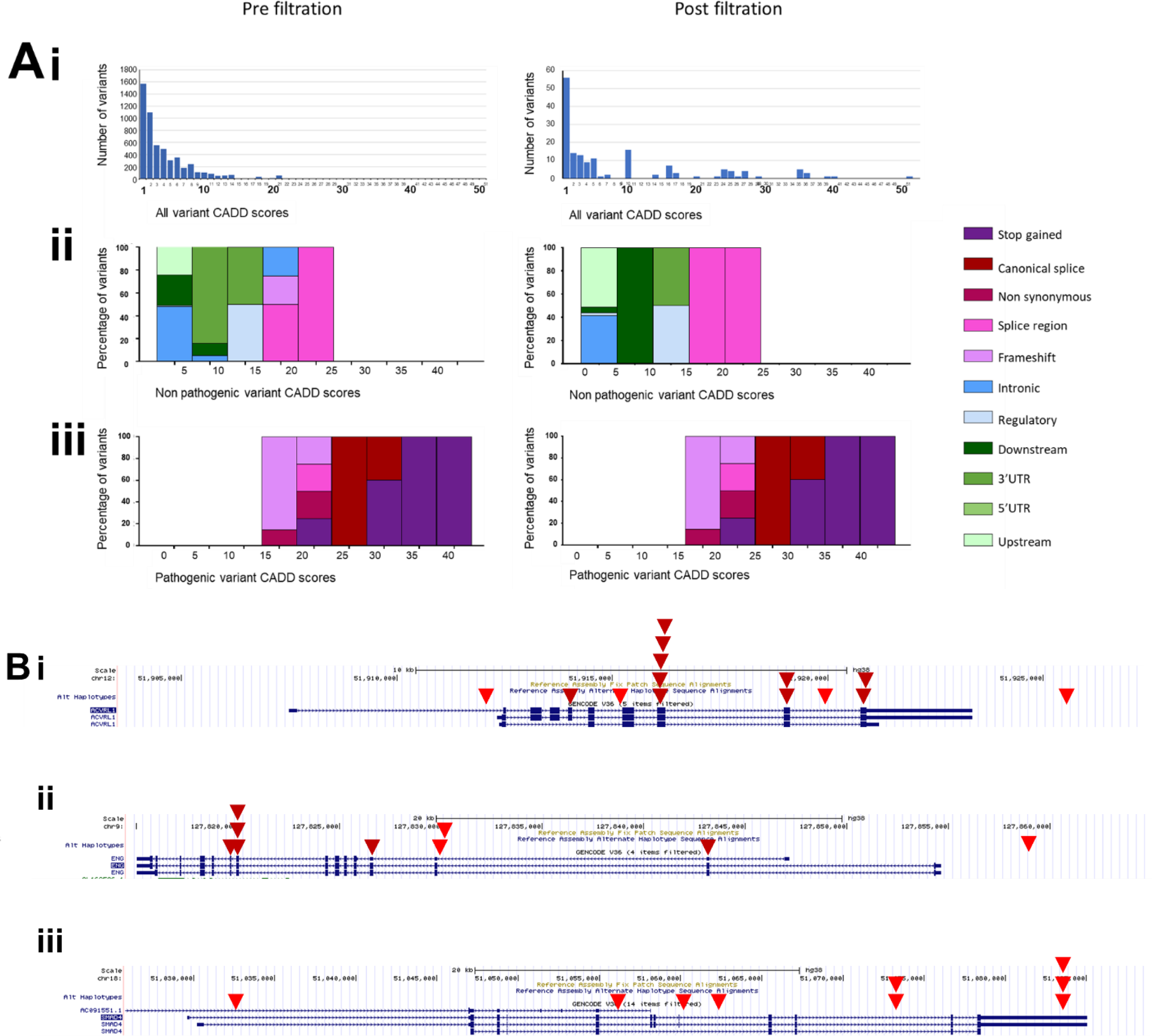
GROFFFY variant-level validation: Comparison of HHT gene variants in the validation and discovery datasets. **A)** Validation dataset: **i)** Total number of variants with indicated CADD scores (note logarithmic scale pre filtration versus linear scale post filtration). **ii)** Non-pathogenic variants by CADD score categories: Molecular subtype are indicated in the key. **iii)** Pathogenic variants by CADD score categories, and molecular subtype as in key. Note identical plots in **iii** pre and post filtration because all pathogenic variants were still present post filtration. **B)** Discovery dataset: Location of GROFFFY-captured variants in the major HHT genes (*10*) **i)** *ACVRL1*, **ii)** *ENG,* **iii)** *SMAD4*. The cartoons include screenshots of GRCh38 from the University of California Santa Cruz (UCSC) Genome Browser (*14,15*), and major transcripts. Red inverted triangles indicate location of variants after application of all filters (dark red for coding/splice regions, bright red for non-coding regions).

### Hot spot of rare deletion variants in the distal *SMAD4* 3’ untranslated region

No coding *SMAD4* variants were identified in the discovery dataset (Fig. 3B). We focused on a hot spot of 3 deletion variants in the 3’ untranslated region (UTR) of *SMAD4* (Fig. 3B). There were two unique variants, one of which was identified in both affected members of a single family. The variants deleted nucleotides 5,519 and 5,649bp distal to the *SMAD4* stop codon, and did not affect any microRNA binding sites (*23*,*24*). The wild-type sequences were consistently expressed in human primary blood outgrowth endothelial cells (BOECs) derived from donors with normal SMAD4 (*25*) (Fig. 4A). General population common variant data also supported the importance of the region: while the 3’ UTR did not contain any expression quantitative trait loci (eQTLs) (*26*) (Fig. 4B), the variants were within the only kilobase of the 3’UTR to contain 3’ alternate polyadenylation QTLs (3’ aQTLs) (*27*) (Fig. 4C).

**Fig. 4:**
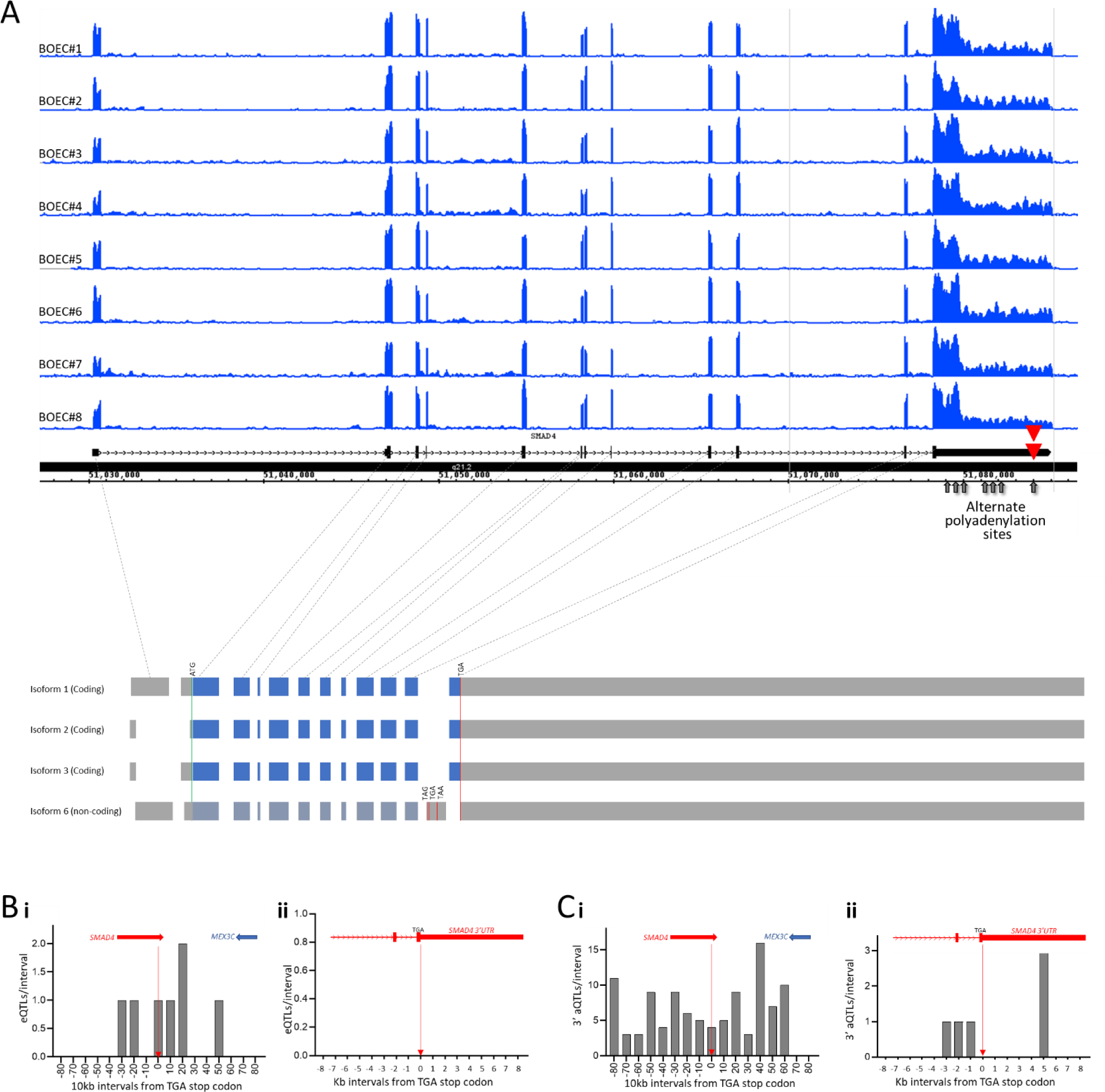
Expression of proto-oncogene *SMAD4* in primary human cells. **A)** Endothelial *SMAD4* total RNA expression: RNASeq data from 8 different cultures of blood outgrowth endothelial cells (BOECs) with normal *SMAD4* sequence (*25*). The consistent peaks sharply define exon boundaries in GRCh38. Sites of start and stop codons, unique filtered variants (red triangles), and the seven alternate cleavage and polyadenylation sites at c.3121, c.3487, c.3791, c.5186, c.5452, c.5615 and c.7709 are highlighted. The cartoon below links the RNASeq expression by grey dotted lines.to the main (upper) and alternate *SMAD4* RefSeq (*28*) splice isoforms that share the final UTR-containing exon, with exons to scale. Blue: coding, grey: non-coding regions. **B)** Number of general population *SMAD4* expression QTLs (eQTLs, (*27*)) per interval of DNA flanking the TGA stop codon, as listed by USCSC CAVIAR tracks (*14,15*) for data from the Genotype Tissue Expression project (GTEx) (*26*). The graphs are centered on the *SMAD4* natural stop codon site (vertical red arrow), with relevant gene loci indicated to scale horizontally above graphs. **i)** Overview of *SMAD4* locus and flanking regions at 10kb intervals. **ii)** Magnified view of penultimate and final exons at 1kb intervals. **C)** Number of general population 3’UTR alternative polyadenylation QTLs (3’aQTLs (*27*)) per kilobase of DNA flanking the TGA stop codon, as determined in GTEx (*26*). **i)** Overview of of *SMAD4* locus and flanking regions at 10kb intervals. **ii)** Magnified view of penultimate and final exons at 1kb intervals. *All graphs are also provided at end of manuscript to enhance clarity*.

### Variants delete nucleotides near the final *SMAD4* alternate polyadenylation site

The *SMAD4* UTR used by all coding transcripts contains 7 alternate polyadenylation site (PAS) AAUAAA hexamers. These cluster in two proximal groups of 3, before a single final AAUAAA at chr18:51,083,977 (Fig. 4A). This final hexamer lay immediately proximal to the two deletion variants, and as expected, (*29*) was flanked by an upstream AU-rich element suited to binding of proteins in the cleavage and polyadenylation (CPA) complex, and downstream repeat elements predicting intermolecular interactions in single stranded RNA that would generate secondary structures to block the progress of RNA polymerase II (Fig. 5A). One GROFFFY-filtered variant deleted the 16 nucleotides sited +3 to +18 from the PAS hexamer with 5 further single nucleotide substitutions, and the second deleted 4 nucleotides in the downstream repetitive element region (Fig. 5B, *Table S6*).

**Fig. 5:**
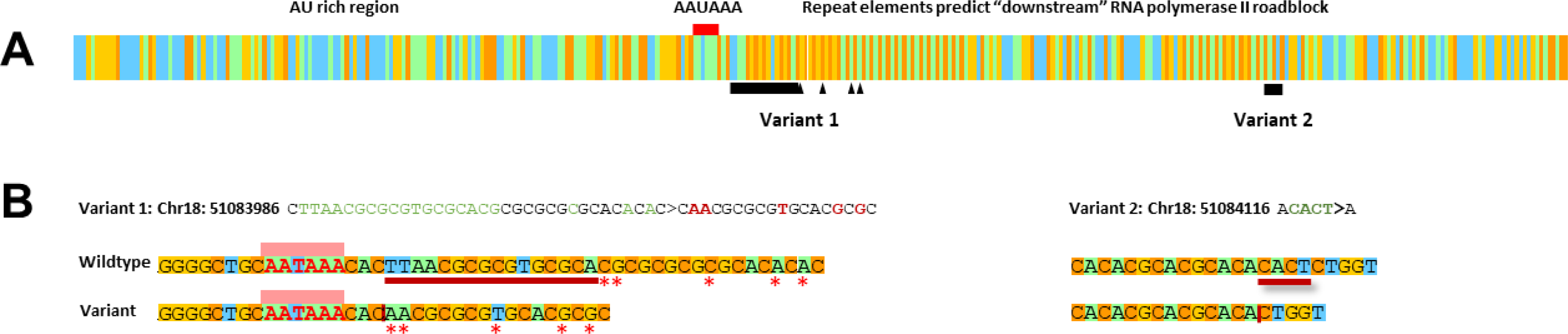
Schematic of *SMAD4* 3’ UTR variants in the context of RNA function. **A)** Color-coded nucleotides 7561_7920 of the *SMAD4* main coding transcript NM_005359. These span the final AAUAAA hexamer (red bar) and include the upstream AU-rich (blue/green) region, downstream repetitive elements, and sites of the two variants. Deleted residues are indicated by black bars, missense substitutions as black triangles. For FASTA format sequences, see *Table S6*. **B)** Variant 1 (chr18:51083986 CTTAACGCGCGTGCGCACGCGCGCGCGCACA>CAACGCGCGTGCACGCG and Variant 2 (chr18:51084116 ACACT>A) in detail. The AAUAAA hexamer is shown by DNA sequence (AATAAA) and highlighted by a pink box, deleted residues by red underline (wildtype) or vertical red line (variants), and missense substitutions as red stars. *Table S6* provides further sequence details.

### The deletion variants disrupt RNA secondary structures required for cleavage and polyadenylation

iFoldRNA secondary structures (*30,31*) visualized using Mol* Viewer (*32*) via the Research Collaboratory for Structural Bioinformatics Protein Data Bank server (*33*), indicated that both deletion variants disturbed secondary structures that substantially altered the sequence context for CPA activity. In wildtype sequence, the AAUAAA hexamer was in a near-linear conformation with stacked pyrimidine and purine rings evident on magnified views (Fig. 6Ai). Strikingly, with the neighboring complex deletion variant, the AAUAAA nucleotides acquired new inter-molecular interactions, lost the stacked alignment of bases, and were incorporated into inaccessible secondary structures (Fig. 6Aii). In contrast, the second variant which deleted 4 nucleotides 134bp downstream of the AAUAAA hexamer, disrupted and unfolded the downstream structured region expected to be the major RNA polymerase II roadblock (*29*) (Fig. 6B).

**Fig. 6:**
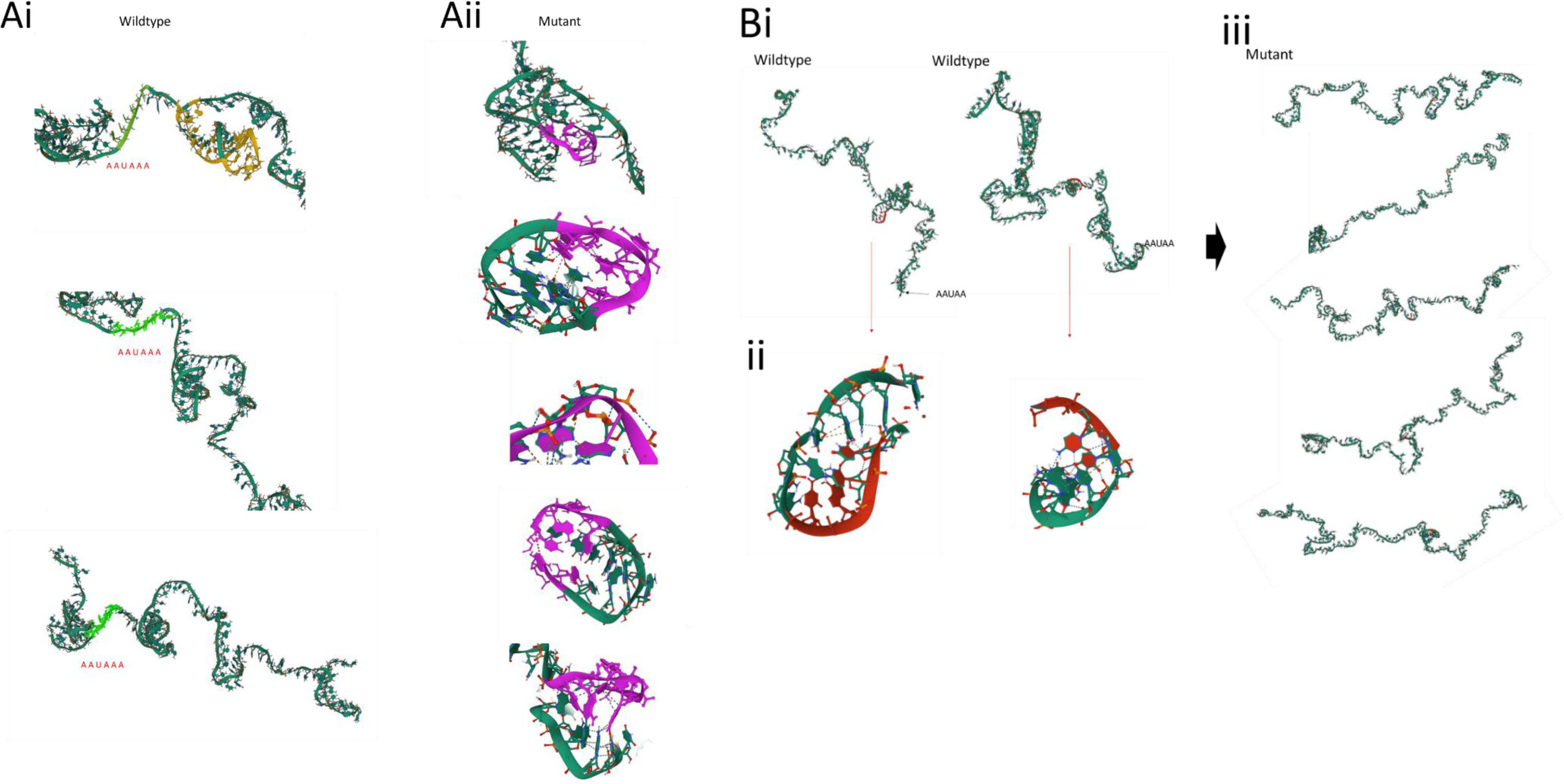
Replicate iFoldRNA structures. iFoldRNA (*30,31*) simulations as visualized in Mol* Viewer (*32,33*). **A)** Variant 1: **i)** Three representative simulations of the wildtype 150 nucleotides spanning the AAUAAA hexamer (light green). The site of the 32 nucleotide deletion/insertion is highlighted in yellow within the upper structure. **ii)** Magnified view of five separate simulations of Variant 1 sequence with AAUAAA hexamer site now highlighted in purple. All 5 simulations were consistent. **B)** Variant 2**: i/ii)** Two representative simulations of wildtype sequence. The site of the 4 nucleotides deleted in the variant are highlighted in red in panoramic (**i**) and magnified (**ii**) views. **iii)** Five simulations of the variant sequence.

### Clinical correlations

All three patients with Variants 1 and 2 had clinically-confirmed HHT (*10-12,34-39*). The first-degree relatives with Variant 2 had no other identified cause to HHT. They each experienced daily nosebleeds, had classical HHT telangiectasia, and one had pulmonary arteriovenous malformations requiring treatment, and hemihypertrophy (left-right axis defect). Gastrointestinal and aortopathy screening had not been considered. The patient with Variant 1 did have a missense variant in *ACVRL1,* though in addition to severe nosebleeds needing blood transfusion and intravenous iron, classical HHT telangiectasia and pulmonary arteriovenous malformations, they had extensive colonic and rectal polyps requiring excision.

### Patient-derived monocyte *SMAD4* RNA Expression

As described in detail in the Appendix, patient and control monocytes were isolated and cultured in conditions predicted to modify 3’UTR use, before RNA sequencing. DESeq2 (*40*) analyses of PolyA-selected RNAs indicated that *SMAD4* polyadenylated transcripts increased after a 1hr hypothermic stress, and this was also seen in 2 patients (*Fig. S3*). However, for the rRNA-depleted libraries representing “total” RNA, variability between control samples assessed by initial DESeq2 analyses was high (*Fig.S3*). This reduced after normalising with low GINI coefficient genes (*41,42*, *Fig. S4*).

Whether normalised to read counts per library, or GINI genes (*41–43*), total *SMAD4* RNA expression was lower in the “inaccessible AAUAAA” Variant 1 donor than 3 normal controls in all conditions (Fig 7Ai). Decrements were also apparent restricting to untreated monocytes, examining exon regions by DEXSEQ (*44*) (Fig 7Bi). In controls, *SMAD4* transcript expression was modified following 1hr cycloheximide 100μg/mL, with lower use of exon region (ER)60 containing the AAUAAA site and variants, consistent with shorter 3’UTRs after stress (*Fig S5, Fig S6*). Despite this, ER60 use was further reduced in the Variant 1 donor after CHX (Fig 7Ci, Fig 7Di) with increased use of penultimate exon regions ER52-55 (Fig 7Di, Fig 7Ei), supporting different 1hr changes in RNA splicing on stress.

**Figure 7.**
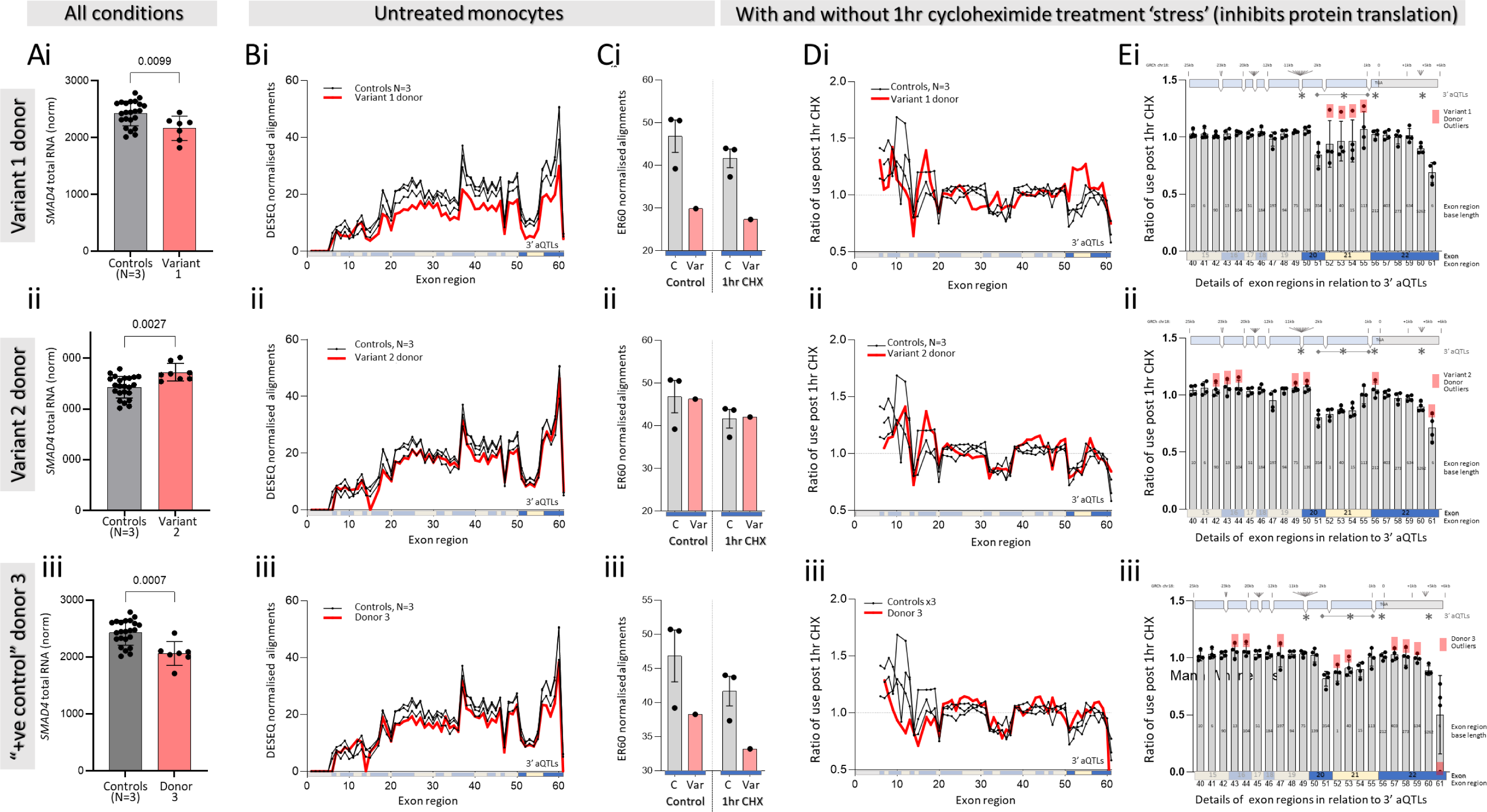
Monocyte *SMAD4* RNA expression in ribosomal (r)RNA-depleted libraries from 3 controls compared to 3 individual patients: (**i)** Variant 1, **ii)** Variant 2, **iii)** an unsolved clinical *SMAD4* positive control. For preceding methodological data on the rRNA depleted libraries, see *Figs. S3-S6*. Monocyte RNA was examined with and without 3 different 1hr stresses (*Fig. S3): SMAD4* splice site changes met DEXSeq2 (*44*) significance only after cycloheximide (CHX). **A)** Total *SMAD4* RNA from control (grey, N=23) and patient donors (red: **i)** Variant 1 N=7; **ii)** Variant 2 N=8; **iii)** Donor 3, N=7) following DESeq2 normalisation (*42,43*) using GINI housekeeper genes (*41,42*). Note contrast between **i/ii** (Variant 1 and 2 donors), but similarities between **i/iii** (Variant 1 and Donor 3)**. B-E)** DEXSEQ (*44*) splicing patterns across 61 exon regions in 22 *SMAD4* exons. **B)** Exon region (ER) use in untreated monocytes by donor, plotting data from the individual patients (red) and the same 3 controls (black). Exons are colour-coded to highlight 3’ aQTL loci. **C)** Use of ER60, the variant-containing 3’UTR region in untreated and CHX-treated monocytes: note again **i/iii** similarities. **D)** The ratio of exon region use between CHX-treated and untreated monocytes, plotted as in **B). E)** The ratios in the final 8 exons (ER40-ER61) containing all ERs differentially used by the Variant 1 and 2 donors after CHX. Each graph is annotated with the genomic DNA origins and kb markers (upper bar); sites of the 3’ aQTLs (*, see Fig 4C); and variant outlier values (red) that were not accompanied by increased polyadenylated transcripts (*Fig S3*).

Total *SMAD4* RNA was higher in the “roadblock unfolding” Variant 2 donor than 3 normal controls across all conditions (Fig 7Aii). Although exon region use was similar to controls in untreated monocytes (Fig 7Bii, Fig Cii), after 1hr cycloheximide, compared to controls there was higher use of regions corresponding to two of the 3’ aQTLs (Fig 7Dii, Eii). We concluded that the contrasting overall expression patterns were consistent with the opposing predictions following RNA modelling of Variants 1 and 2; that Variant 2 data also supported different 1hr changes in RNA splicing after CHX stress, but that precise transcript changes would need to be the subject of future RNA studies.

### Validation of positive control variant

The third donor had been recruited as a SMAD4 positive control due to HHT-JP syndrome (colonic and gastric polyposis; HHT nosebleeds; HHT mucocutaneous telangiectasia, pulmonary AVMs treated by embolization, and antecedent HHT-JP family history). However, no *SMAD4* variant had been identified by clinical service panel testing, the 100,000 Genomes Project clinical pipelines, or by GROFFFY. Total monocyte *SMAD4* expression levels were lower than controls (Fig 7Aiii), and similar to Variant 1 (Fig 7Ai/iii) with additional similarities to Variants 1 and 2 post cycloheximide (Fig 7E). A new team member blinded to the findings and project, was invited to examine the raw *SMAD4* WGS data in the binary alignment map (bam) file and identified a single exonic variant in the donor’s DNA (*Fig. S7*). This was sited between Variants 1 and 2 in the 3’UTR, with the complex insertion/rearrangement separated from Variant 1 by only two bases (*Fig. S7*).

## DISCUSSION

We have presented and validated a system that synthesizes biologically-generated signals of function in order to filter out variants in DNA regions with no such evidence of functionality. This generically applicable method was highly effective in reducing the number of WGS variants from almost 5 million per individual to an average of ∼21,000. Critically, the method retained pathogenic variants already known in a validation dataset, and identified ultra-rare, disease-associated variants in the distal *SMAD4* 3’ UTR. These variants disrupted RNA secondary structures required for cleavage and polyadenylation that is often overlooked as an essential pre-requisite for protein translation, and subsequent RNASeq and clinical correlations supported *SMAD4* etiology.

Study strengths include the development and application of an unbiased, genome-wide method with no prior assumptions. Of other variant filtration methods already used in WGS, most depend on union and intersection rules of existing annotation tracks. The candidate cis-regulatory elements file produced by ENCODE has been particularly favored with its specific predictions of each possible CRE position and size. By using the raw biological data providing broader areas for inclusion, GROFFFY may better suit the purpose of a first pass filter for definition of variants worthy of further study, than computational predicted files with potential false negatives. Simultaneous evaluation of nearly 100 patients with a similar phenotype enabled resource direction to unstudied non-coding sequences where multiple rare, high impact variants were identified. Study strength was further augmented by replicate RNASeq expression data from primary human endothelial cells, the cell type responsible for the *SMAD4* clinical phenotype (HHT) where causal loss-of-function variants were being sought, and Genomics England clinician contact pathways that identified patients and *SMAD4-*specific phenotypes after the draft manuscript was approved for submission. This also enabled patient recontact, providing evidence from patient-derived monocytes to support perturbations in *SMAD4* RNA expression. Additionally, extensive open-source datasets and code enabled exploration of common human variation responsible for *SMAD4* QTLs impacted by the identified variants, while the variants themselves highlighted an emerging field in biology that has had limited recognition in medicine.

A potential study weakness, the presented discovery elements that focus on a single gene, is justified because of the immediate pathway to translational impact. In addition to somatic cancer genetic diagnostics, early diagnosis of a germline heterozygous *SMAD4* loss-of-function allele offers proven methods to save lives and emergency healthcare resources by institution of gastrointestinal (from adolescence) and aortic screens (*9*), in addition to standard HHT screening and pre-symptomatic interventions (*11,12*). While detailed mechanistic dissection can be the subject of future work, the presented data already support immediate extension of the *SMAD4* regions included in biological and virtual gene panels for patients with HHT, juvenile polyposis and cancer to include the 3’UTR sequences flanking the final AAUAAA hexamer. For the HHT patients harboring the identified variants, there seems sufficient evidence for them to be considered as “likely *SMAD4* HHT” for at least one round of endoscopic and echocardiographic surveillance, while functional studies are pending. For other HHT patients where conventional screening of HHT genes has not identified a causal variant, the possibility of undetected *SMAD4* variation can be considered.

Alternate polyadenylation has not been explored to date for *SMAD4*, or for other heritable diseases beyond triplet expansion neurodegenerative diseases (*29,45*). Long 3’UTRs with their abundance of regulatory motifs provide greater opportunity for regulatory control than short 3’ UTRs, while switching between alternate polyadenylation sites to provide shorter or longer 3’UTRs is increasingly recognized to modify protein translation, for example differentially transporting mRNAs to condensates which can result in translation repression or enrichment in specified cellular regions or states (*29*). Our data suggest this is particularly important for regulation of *SMAD4*, a ubiquitous and essential protein with diverse functions (*8,9*) where ∼7kb of 3’ UTR is transcribed at high levels in coding and non-coding transcripts (Fig. 4, Fig. 7, *Fig. S5*, *Fig. S6*). Recent data highlight that polyadenylation sites differ in strength: weaker proximal CPA sites are used in genes with cell type-specific transcription, (requiring transcriptional enhancers to strengthen CPA activity), while distal and single PAS sites are strongest to ensure mature mRNAs are produced (*46*).

As recently reviewed (*29*), cleavage and polyadenylation occurs while RNA polymerase II (Pol II) is transcribing a gene, and is regulated by Pol II elongation dynamics. Pol II pausing immediately downstream to a final AAUAAA hexamer CPA cleavage site is necessary in order to enable CPA complex assembly and co-transcriptional addition of the “poly-A tail” that is essential for mRNA generation and subsequent protein translation (Fig. 5). If at the final polyadenylation site, the full cycle of polymerase pausing, CPA complex binding and cleavage/polyadenylation is impaired, different sites and efficiency of polyadenylation would modify function. Our current data examining 1hr stress responses when the cell has to rely predominantly on reuse of existing RNA transcripts, highlight further novel mechanisms to explore. These include 3’ UTR variant impacts on alternate splice site selection, and maintenance of polyadenylated transcripts that may be less successfully achieved in the setting of stress conditions necessitating rapid changes (*Fig S3A*). The potential to facilitate future development of 3’UTR therapeutics is augmented given repetitive regions of pol II “roadblocks” provide fertile and previously hidden substrates for impactful human DNA variation,

In conclusion, we present and validate a filter that reduces the overwhelming number of variants identified by WGS, while retaining functional genome variation of importance to patients. Exposure of non-coding variants in the top 10 percentile of deleteriousness, and clusters in unexplored genomic regions, enhances the near-term value of WGS. The GROFFFY filter enabled identification of rare *SMAD4* variants that disrupt the final site for RNA cleavage and polyadenylation, necessary for protein production. However, the full extent to which rare **<u>s</u>**tress **i**mpact, **f**unctional **<u>a</u>**lternate **<u>p</u>**olyadenylation **<u>s</u>**ite (SIFAPS) variants contribute to diseases will only be exposed if untranslated sequences spanning the sites are included in virtual and physical diagnostic gene panels. Wider use of WGS, or inclusion of 3’aQTL UTR regions in exome-based sequencing is recommended to capture relevant patient-specific variants.

## MATERIALS AND METHODS

### Study Design

The main elements of Study Design are outlined in Fig. 8. Following recruitment of patients with hereditary haemorrhagic telangiectasia (HHT) to the 100,000 Genomes Project (black arrows), the GROFFFY filter was designed as indicated in colored boxes, and applied to the WGS data files from the 100,000 Genomes Project.

**Fig. 8.**
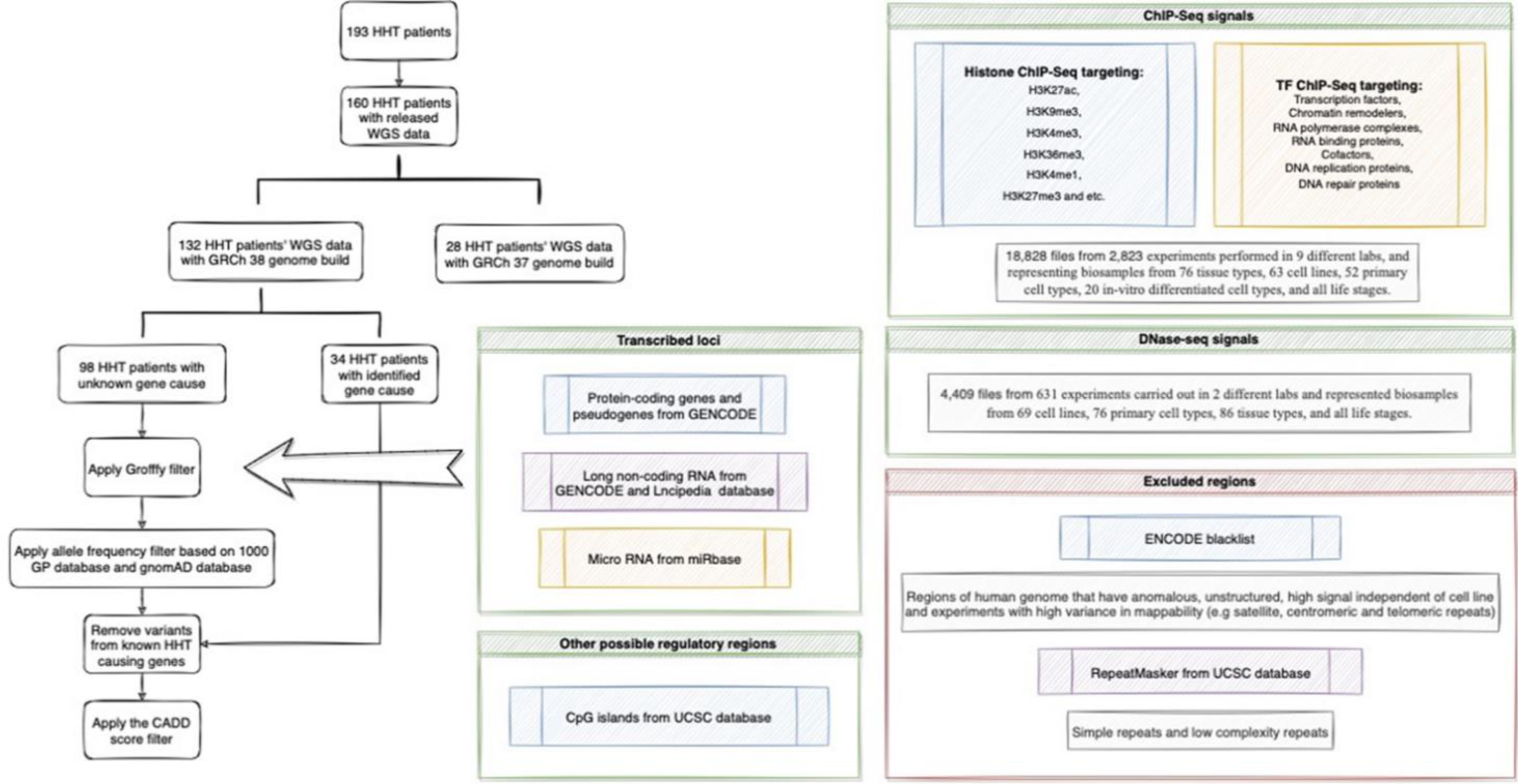
GROFFFY Study Protocol: Flow chart illustrating sequence of stages described in the text and online Data Supplement

### Patient recruitment and sequencing

The 100,000 Genomes Project was set up by the UK Department of Health and Social Security in 2013, to sequence whole genomes from National Health Service (NHS) patients. The study received ethical approval from the Health Research Authority (HRA) Committee East England-Cambridge South (REC Ref 14/EE/1112), and all participants provided written consent. Anonymized raw sequencing data were available in the Genomics England Research Environment (*7,47*). Separately, in a clinical diagnostic pipeline, Genomics England performed data alignments and variant classifications fed back to recruiting clinicians.

The cohort recruited with HHT were particularly suited for *GROFFFY* methodological validation processes because a subset had not undergone prior genetic testing, and because clinical pipelines were incomplete at the time of *GROFFFY* analyses. This resulted in a validation dataset of 34 WGS sequences where clinical pipelines had identified a causal variant (*35–39*), and discovery dataset of 98 WGS sequences where some DNAs were expected to have heterozygous loss-of-function variants in *ACVRL1*, *ENG* or *SMAD4* (*10*).

### Generating GROFFFY

Genomic coordinates of regions included in GROFFFY were generated from publicly available databases using the Imperial College High Performing Computing service. Experimentally-derived biological data were used in preference to computational predicted files with potential for false negatives. Genomic coordinates were extracted from data aligned to GRCh38 (*13*), and excluded data originating in cancer cells. Regions being selected for are described in *Tables S1-S4* which provide full details of coordinate derivation from transcribed loci and candidate regulatory element (cRE) regions (*6,17,18*). Following merging of 18,828 bed files from 3,454 experiments (*5,6*), sequences in the ENCODE blacklist (*48*) and RepeatMasker (*49*) were excluded. In detail:

***Genomic coordinates for candidate regulatory element (cRE) regions*** were generated using data from the ENCODE Encyclopedia registry which includes data from both ENCODE (*4,5,50*) and the NIH Roadmap Epigenomics Consortia (*51*). We downloaded the call sets itemized in *Table S3* and *Table S4* from the ENCODE portal (*4,5*). By merging DNA binding call sets (*50*) and representative DNase hypersensitivity site (rDHS) call sets, (*52*) a rough prediction of all cREs was made. Only data aligned to GRCh38 were retained. Data generated from cancer cells were excluded as cancer cells’ genome are usually heavily modified and rearranged (*53*).

a) For DNA binding data, Histone ChIP-seq and Transcription Factor ChIP-seq which target histones, transcription factors, chromatin remodelers, RNA polymerase complexes, RNA binding proteins, cofactors, DNA replication proteins and DNA repair proteins, were searched. We downloaded the call sets from the ENCODE portal (*4*) as indicated in *Table S3*. The 18,828 downloaded files (*Table S3*) were from 2,823 experiments performed in 9 different labs, and representing biosamples from 76 tissue types, 63 cell lines, 52 primary cell types, 20 in-vitro differentiated cell types, and all life stages. Downloaded bed files were divided into 10 subgroups with the first 9 subgroups containing 2,000 bed files and the last subgroup containing 828 bed files. Bed files from each subgroup were merged together using BEDOPS (*54*) and then merged bed files from each subgroup were joined together last to obtain all DNA binding regions. The merged bed file for DNA-binding regions was 49,941,695 Kb before sorting.
b) For rDHSs, DNA accessibility experiments were searched in the ENCODE Encyclopedia database (*5*). We downloaded 4,409 files as indicated in *Table S4*. These were from 631 experiments carried out in 2 different labs and represented biosamples from 69 cell lines, 76 primary cell types, 86 tissue types, and all life stages. Bed files were merged together directly using BEDOPS (V2.4.26) (*54*) to obtain all accessible DNA regions.
c) Region coordinates for CpG islands were downloaded from the UCSC database (*14,15*) using Rsync command tool. The downloaded file was in txt format and was converted to bed files using awk function in the Linux system (*55*). The converted bed file was then sorted by location using BEDOPS (*54*).

***Genomic coordinates for transcribed loci*** were extracted as follows:

d) GENCODE human genome annotation version 31 (*16*) for GRCh38 (*13*) was downloaded from the UCSC database (*14,15*) using Rsync in the command line. All gene coordinates were extracted by using awk function (including protein-coding gene and pseudogenes). The downloaded file was in gff3 format and was converted to bed files using BEDOPS (*54*).
e) Long non-coding RNA annotations (lncRNA) were downloaded from both GENCODE (release 31) (*16*) and lncipedia database (version 5.2) (*17*). LncRNA coordinates were merged together using BEDOPS (*54*) to obtain all possible lncRNA gene regions.
f) Micro RNA (miRNA) annotations were downloaded from miRbase (release 22.1) (*18*). The downloaded file was in gff3 format and was converted to bed files using BEDOPS (*54*).

***Genomic coordinates of regions excluded*** were identified from the ENCODE blacklist which was downloaded directly from the ENCODE (*56*) project website in bed format, and RepeatMasker (*57*) which was downloaded from the UCSC database (*14,15*) in txt format. The Linux command awk was used to grep out region coordinates, and BEDOPS (*54*) was used to sort the file.

***To assimilate for GROFFFY***, separately, the Ubuntu shell (version 16.04.2 LTS based on Linux 4.4.0-64-generic x86_64 system) was launched for the Genomics England Research Environment (*47*), where the final genomics coordinates for GROFFFY were transferred. WGS variant data were examined after analysis by the Illumina WGS Service Informatics pipeline. This used Illumina Issac (*58*) for sequence alignment, and Starling (*59*) to identify variants. The output files were in vcf files with .gz compression and decompressed using Gzip (Version 1.6) (*60*). Pandas module version 0.22.0 (*61*) in Python (version 3.6.5) was used to process vcf files. Pegasus, the High-Performance computer cluster of Genomics England, was used to run computationally intensive jobs, submitted to the Load Sharing Facility (LSF).

The Intersect function of Bedtools version 2.26.0 (*62*) was then used to identify WGS variants from vcf files that were in the GROFFFY bed file regions. Option -header was used to remove any headers, and option -wa was used to ensure the output file format was the same as the input vcf file. For the Intersect function, any intersection with GROFFFY was outputed to result files, even if some part of the variation was outside of the filter region. Annotations of the WGS vcf files were carried out using the Ensembl Variant Effect Predictor (VEP) version 96.3 (*63*), based on Perl version 5.24 (*64*), SAMtools version 1.5 (*65*) specifically SAMtools HTSlib version 1.5 (*65*), and a list of options to optimize the process (*Table S2).* A Python script was written to produce 66 shell scripts where each shell script contained 2 annotation jobs. R version 3.5.1 (*66*), within R studio version 13.4.0 (*67*) downloaded from the Comprehensive R Archive Network (*68*), was used to perform statistical tests. Paired datasets were analysed using the non-parametric Wilcoxon rank-sum test; multiple datasets by the Kruskal-Wallis rank-sum test and Dunn’s multiple comparison test.

Regions from the ENCODE Blacklist (*48*), and RepeatMasker (*49*) were subtracted from regions selected for using the ‘difference’ option in BEDOPS (*54*). All bed files were merged together to obtain the selected genomic regions. Numeric data and Python scripts were approved for export through the Research Environment AirLock under subproject RR42 (HHT-Gene-Stop, *Table S7*).

### GROFFFY analysis of whole genome sequencing data

Stepwise filters excluded variants where general population allele frequency exceeded 0.0002 in the 1000 Genome Project (*20*) or gnomAD (*21*) databases; synonymous variants not in splice regions; all non HHT-causal variants in the Validation Set HHT DNAs; and variants with a CADD score <10 (*22*). In detail:

***An autosomal dominant-specific disease application step*** was included as a high stringency “white list” filter. For this, the annotated WGS files were retrieved for the 34 Validation Set DNAs where a causative variant had already been identified in known HHT genes through clinical pipelines (*35–38, 69*). Variant information was collected through unique variant IDs consisting of chromosome number, variant starting position, reference sequence and altered sequence (e.g. chr1:111_C/TTT). To confirm that no two variants were represented by the same variant ID, the full list was compared to a set where only unique values were stored, and shown to be identical. The variant IDs were integrated in a white-list. Exclusion of these white-listed variants in other patients was performed using the isin function of Pandas (*61*) module: Any variant in the white list was deleted from the vcf files of the target set DNAs, and the number of variants after exclusion was recorded and outputed to txt files.

***For CADD score filtration and prioritization,*** the plugin option of VEP was used to annotate variants with CADD scores (*22*) in the enclosed Research Environment: databases for SNV annotation (version 1.5) and small indel annotation (version 1.5) which were pre-installed in the Research Environment were indexed. As the annotation of CADD score was quite slow, the process was put towards the end of the analysis pipeline, so that there were fewer variants that needed to be annotated. Prioritization by CADD score was performed by generating further customized Python scripts. The CADD PHRED-scale score for all 9 billion SNVs and millions of small indels was extracted from the information column. Variants absent from the CADD score database were represented by an empty string by default and were replaced by number 999 instead. Variants with a PHRED score less than 10 were removed so that both variants with top 10 percentiles deleteriousness and variants absent from the database were prioritized. The processed files were stored in vcf format.

### Export of variant coordinates and bioinformatic analyses

Following approval for export through AirLock (*Table S7*), variant genomic coordinates were visualized in the UCSC Genome Browser (*14,15*). Endothelial expression of *SMAD4* was examined in whole transcriptome data from primary human BOECs (*25*). Binary sequence alignment map (bam) files aligned to GRCh38 (*13*) were analyzed in Galaxy Version 2.4.1 (*70*) and the Integrated Genome Browser (IGB) 9.1.8 (*71*).

3’UTR alternative polyadenylation quantitative trait loci (3’aQTLs) from 46 tissues isolated from 467 individuals in GTEx (*26*) were sourced through the 3’aQTL Atlas (*27*). Genetic variants likely affecting gene expression in GTEx V8 (*26*) data release were captured from UCSC (*14,15*) CAVIAR tracks, which define high confidence gene expression QTLs within 1MB of gene transcription start sites (cis-eQTLs) (*14,15*).

All variants were independently verified by Genomics England. Impact on microRNA binding sites was examined through TargetScan Human Release 8.0 (*23*) and miRDB (*24*). RNA structure predictions were performed using iFoldRNA v2.0 (*30,31*) without restraints, and final models were visualized using Mol* Viewer (*32*) via the Research Collaboratory for Structural Bioinformatics Protein Data Bank server (*33*).

### Clinical re-contact, correlations, and re-sampling

Genomics England “Contact the Clinician” forms were submitted through the Research Environment and clinicians who had recruited the patients were contacted and joined the research team. Clinical correlations were performed through North Thames and South-West NHS Genomic Medicine Service Alliances. Patients were contacted by their clinicians and provided written consent for publication after reviewing the relevant sections of the manuscript.

Patients also consented to further blood samples together with 3 healthy volunteers and a further unsolved patient recruited to the 100,000 Genomes Project with a *SMAD4*^+/-^ phenotype. This study was approved by the East of Scotland Research Ethics Service (EoSRES: 16/ES/0095), and the 6 participants provided written informed consent. Using methods we have developed to perform experimental treatments on human cells while resuspended in endogenous plasma (*72–74*), peripheral blood mononuclear cells (PBMCs, ‘monocytes’) were prepared using BD Vacutainer® CPT™ tubes (Bunzl Healthcare, Coalville, UK) according to manufacturer’s instructions with minor modifications. As detailed further in the Appendix, these were to provide comparative resources of cells in stressed and unstressed states, where alternate transcripts/exon region use could potentially be impacted by modified efficiency of final AAUAAA cleavage and polyadenylation due to the 3’UTR variants.

Briefly, immediately after venesection, the blood was gently remixed by inverting 8-10 times, and centrifuged within 2 hours of collection for 30 minutes at 1600 relative centrifugal force (RCF) at room temperature. Monocytes and plasma were collected by pipetting from above the gel layer, transferred to a single 50ml tube for each donor, and gently inverted to resuspend. After monocyte resuspension in plasma, for each donor, equal volumes were distributed to separate experimental treatment tubes, prewarmed at 37°C for 10 minutes, then subjected to 4 different treatment conditions for 1 hour including control at 37°C, and low temperature in a 32°C waterbath for 1hr to mimic the stress incurred at the threshold between mild and moderate hypothermia (*75*). Additional stresses previously optimized in our laboratory (*72-74,76,77*) were inhibition of translation by cycloheximide 100μg/ml (cycloheximide inhibits eukaryotic translation elongation by mechanisms include binding to the 60S ribosomal subunit E-site (*78,79*)) and a clinically-relevant mild reactive oxygen species (ROS) stress using ferric citrate 10μmol/L (*77,80*). After 1hr, all tubes were centrifuged at 520 RCF at room temperature for 15 minutes. Cell pellets were lysed in Tri reagent (Cambridge Bioscience Ltd, Cambridge, UK) before distribution to replicate tubes for paired rRNA-depleted and polyA-selected RNA sequencing library generation.

### RNA Sequencing and Differential Expression Analyses

RNA extraction and quality control for 96 samples was performed by Genewiz (Leipzig, Germany). For RNASeq library preparations, 48 samples were polyA selected for polyadenylated RNA enrichment, and 48 paired samples underwent ribosomal (r)RNA depletion. RNA was fragmented and random primed for first and second strand cDNA synthesis, end repair, 5’ phosphorylation, dA-tailing, adaptor ligation, PCR enrichment and Illumina HiSeq sequencing using paired-end 150bp reads (Genewiz, Leipzig, Germany). Sequenced reads were trimmed using Trimmomatic v.0.36 (*81*), aligned to Homo sapiens GRCh38 (*13*) using STAR aligner v2.5.2b, and unique gene reads that fell within exon regions counted using Subread package v1.5.2 (Genewiz, Leipzig, Germany).

Blinded to the types of donors and treatments, Genewiz performed differential gene expression analyses using DESeq2 (*40*), and differential exon expression using DEXSeq (*44*) to identify differentially spliced genes by testing for significant differences in read counts on exon regions (and junctions) of the genes. In DEXSeq (*44*), read counts are normalised by size factors: Contributions to the average are weighted by the reciprocal of an estimate of their sampling variance, and the expected variance used to derive weights for the “balanced” coefficients reported as estimates for the strengths of differential exon usage and DEXSeq plotting, that are of similar magnitude to the original read counts (*44*). The output indicates alternative transcript isoform regulation, noting individual exon region assignment is reliable as long as only a small fraction of counting regions (bins) in the gene is called significant (*44*). These conditions were noted to be met for *SMAD4* (*Fig. S7)*, supporting inferences about particular exon regions.

Noting control variability in initial DESeq2 analyses (*Fig. S3*), the least variable of human transcripts (the 25 genes with GINI Coefficients (GCs)<0.15 in diverse cells (*41,42*)) were used to evaluate individual library quality (*Fig. S4*), and subsequently employed for DESeq2 normalisation. For these normalisations, the intra-assay coefficient of variation (CV, *100*standard deviation (SD)/mean*) (*82*) was calculated for replicate pairs using alignment per gene adjusted for total read counts per library, and analyses restricted to libraries where >50% of GINI genes had a CV<10% (‘met CV10’). Three rRNA depletion datasets failed this quality control. The remaining datasets were DeSeq2 normalised (*44*) using the GINI genes as housekeepers, when the ratio of alignment counts for each selected housekeeper gene in each dataset to the geometric mean of that gene was calculated across the remaining 45 datasets from rRNA-depleted libraries. The median value of these ratios in each library was used to generate the ‘size factor’ to scale that library’s alignments (*Table S8*).

### Statistics

Descriptive statistical analyses were performed in Python and STATA v17.0 (Statacorp, College Station, Texas). Comparative statistics of the number of variants before and after filtration was performed in Python using Mann Whitney two-group comparisons. RNASeq expression was analysed in STATA v17.0 (Statacorp, College Station, Texas) and GraphPad Prism 9 (GraphPad Software, San Diego, CA), compared using Kruskal Wallis and Dunn’s post test applied for selected pairwise comparisons. iFoldRNA v2.0 computational predictions (*30,31*) were performed without restraints.

## Data Availability

The publicly available file accession numbers used to generate the code are provided in full within the Data Supplement and have been submitted to the NCBI BioProject database (https://www.ncbi.nlm.nih.gov/bioproject/) under accession number PRJNA596860, referencing the WGS data source under accession number SAMN13640532. Primary data from the 100,000 Genomes Project, which are held in a secure Research Environment, are available to registered users. Please see https://www.genomicsengland.co.uk/about-gecip/for-gecip-members/data-and-data-access for further information.

## Supplementary Appendix

Figs. S1 to S7

Tables S1 to S8

## Acknowledgments

This research was made possible through access to the data and findings generated by the 100,000 Genomes Project. The 100,000 Genomes Project is managed by Genomics England Limited (a wholly owned company of the Department of Health and Social Care). The 100,000 Genomes Project is funded by the National Institute for Health Research (NIHR) and NHS England. The Wellcome Trust, Cancer Research UK and the Medical Research Council have also funded research infrastructure. The 100,000 Genomes Project uses data provided by patients and collected by the National Health Service as part of their care and support. We thank the National Health Service staff of the UK Genomic Medicine Centres and the participants for their willing participation; the Genomics England Clinical Research Interface team, specifically Susan Walker, for separately reviewing bam file variant sequences; Santiago Vernia and Michael Hubank for helpful discussions; and our academic and public partners within the NIHR Imperial BRC’s Social Genetic and Environmental Determinants of Health (SGE) theme. We specifically thank the presented families for confirmation of their clinical phenotypes and consent to share in this manuscript. The views expressed are those of the authors and not necessarily those of funders, the NHS, the NIHR, or the Department of Health and Social Care.

## Author contributions

SX devised and generated the GROFFFY approach, devised all scripts to generate GROFFFY, and generated all GROFFFY numeric data, Figs. 1, 2 and 8, Figs. S1 and S2, and Tables S1, S2, S3, S4 S5, and S7. ZK advised on Linux and script generation. DM interrogated Donor 3 bam files. DL assisted in monocyte cultures. DP, AB, MBH, and MAA performed BOEC cultures and RNA sequencing. AA designed primers for validations. AM contributed to patient recruitment. SW contributed to clinical correlations. NV advised on *SMAD4* regulation. GERC performed all sequencing. MJC contributed to specific project set up at Genomics England. CLS recruited patients and performed clinical correlations; devised concepts and advised on GROFFFY approaches; devised and performed monocyte cultures; devised and performed in-house endothelial and monocyte RNASeq and variant level data analyses; generated Figs. 3, 4, 5, 6 and 7, Figs. S3, S4, S5, S6 and S7, Tables S6, S7 and S8, and wrote the manuscript. All authors have reviewed and approved the final manuscript.

## The Genomics England Research Consortium Members comprised on 8^th^ May 2022

Ambrose, J. C. 1 ; Arumugam, P.1 ; Bevers, R.1 ; Bleda, M. 1 ; Boardman-Pretty, F. 1,2 ; Boustred, C. R. 1 ; Brittain, H.1 ; Brown, M.A.; Caulfield, M. J.1,2 ; Chan, G. C. 1 ; Giess A. 1; Griffin, J. N. ; Hamblin, A.1; Henderson, S.1,2; Hubbard, T. J. P. 1 ; Jackson, R. 1 ; Jones, L. J. 1,2; Kasperaviciute, D. 1,2 ; Kayikci, M. 1 ; Kousathanas, A. 1; Lahnstein, L. 1 ; Lakey, A.; Leigh, S. E. A. 1 ; Leong, I. U. S. 1 ; Lopez, F. J. 1 ; Maleady-Crowe, F. 1 ; McEntagart, M.1; Minneci F. 1 ; Mitchell, J. 1 ; Moutsianas, L. 1,2 ; Mueller, M. 1,2 ; Murugaesu, N. 1; Need, A. C. 1,2 ; O‘Donovan P. 1; Odhams, C. A. 1 ; Patch, C. 1,2 ; Perez-Gil, D. 1 ; Pereira, M. B.1 ; Pullinger, J. 1 ; Rahim, T. 1 ; Rendon, A. 1 ; Rogers, T. 1 ; Savage, K. 1 ; Sawant, K. 1; Scott, R. H. 1 ; Siddiq, A. 1 ; Sieghart, A. 1 ; Smith, S. C. 1; Sosinsky, A. 1,2 ; Stuckey, A. 1 ; Tanguy M. 1 ; Taylor Tavares, A. L.1; Thomas, E. R. A. 1,2 ; Thompson, S. R. 1 ; Tucci, A. 1,2 ; Welland, M. J. 1 ; Williams, E. 1 ; Witkowska, K. 1,2 ; Wood, S. M. 1,2; Zarowiecki, M. 1 .

1. Genomics England, London, UK

2. William Harvey Research Institute, Queen Mary University of London, London, EC1M 6BQ, UK.

