## Supplementary Appendix for "Functional filter for whole genome sequence data identifies stress impact, non-coding alternate polyadenylation site variants >5kb from coding DNA"

[1]National Heart and Lung Institute, Imperial College London; London, UK. [2] NIHR Imperial Biomedical Research Centre; London, UK. [3]Topgen Biopharm Technology Co. Ltd; Shanghai, China.[4] Women's, Children's & Clinical Support (Pharmacy), Imperial College Healthcare NHS Trust; London, UK [5] School of Molecular and Cellular Medicine, University of Bristol; Bristol UK. [6] Indiana University School of Medicine; Indianapolis, USA.[7] School of Medicine, Medical Sciences and Nutrition, University of Aberdeen; Aberdeen, UK.[8] William Harvey Research Institute, Queen Mary University of London: London, UK. [9] Genomics England: London, UK, [10] Specialist Medicine, Imperial College Healthcare NHS Trust; London, UK

† Current address: Big Data Institute, University of Oxford, Oxford, UK.‡ A full list of these authors is given below.

#### SUPPLEMENTAL APPENDIX

The Genomics England Research Consortium Members 8th May 2022

Ambrose, J. C. 1 ; Arumugam, P.1 ; Bevers, R.1 ; Bleda, M. 1 ; Boardman-Pretty, F. 1,2 ; Boustred, C. R. 1 ; Brittain, H.1 ; Brown, M.A.; Caulfield, M. J.1,2 ; Chan, G. C. 1 ; Giess A. 1; Griffin, J. N. ; Hamblin, A.1; Henderson, S.1,2; Hubbard, T. J. P. 1 ; Jackson, R. 1 ; Jones, L. J. 1,2; Kasperaviciute, D. 1,2 ; Kayikci, M. 1 ; Kousathanas, A. 1; Lahnstein, L. 1 ; Lakey, A.; Leigh, S. E. A. 1 ; Leong, I. U. S. 1 ; Lopez, F. J. 1 ; Maleady-Crowe, F. 1 ; McEntagart, M.1; Minneci F. 1 ; Mitchell, J. 1 ; Moutsianas, L. 1,2 ; Mueller, M. 1,2 ; Murugaesu, N. 1; Need, A. C. 1,2 ; O'Donovan P. 1; Odhams, C. A. 1 ; Patch, C. 1,2 ; Perez-Gil, D. 1 ; Pereira, M. B.1 ; Pullinger, J. 1 ; Rahim, T. 1 ; Rendon, A. 1 ; Rogers, T. 1 ; Savage, K. 1 ; Sawant, K. 1; Scott, R. H. 1 ; Siddiq, A. 1 ; Sieghart, A. 1 ; Smith, S. C. 1; Sosinsky, A. 1,2 ; Stuckey, A. 1 ; Tanguy M. 1 ; Taylor Tavares, A. L.1; Thomas, E. R. A. 1,2 ; Thompson, S. R. 1 ; Tucci, A. 1,2 ; Welland, M. J. 1 ; Williams, E. 1 ; Witkowska, K. 1,2 ; Wood, S. M. 1,2; Zarowiecki, M. 1 .

1. Genomics England, London, UK
2. William Harvey Research Institute, Queen Mary University of London, London, EC1M 6BQ, UK.

### SUPPLEMENTARY METHODS:

#### Details of monocyte sampling

Peripheral blood mononuclear cells (PBMCs, 'monocytes') were prepared according to manufacturer's instructions using BD Vacutainer® CPT™ tubes (Bunzl Healthcare, Coalville, UK). After resuspension in endogenous plasma, equal volumes were distributed to separate experimental treatment tubes placed at 37°C to prewarm for 10 minutes, before simultaneous 1hr treatments to provide four states (control; hypothermia; translation inhibition; reactive oxygen) with likely alternate 3' untranslated region use[1]. Two sets were treated with 1/1000 Dulbeccos Modified Eagle Medium (DMEM) supplemented with 10% fetal calf serum (FCS): one was the control incubated in a 37°C, 5% $CO_2$ incubator, with the other having a 1hr hypothermic stress in a 32°C waterbath to mimic the threshold between mild and moderate hypothermia[2]. Before their 1hr incubation at 37°C, 5% $CO_2$ the third set of tubes were treated with 10mg/ml cycloheximide[3-7] to a final concentration of 100µg/ml, and fourth with 10mM filter-sterilised and freshly prepared ferric citrate to a final concentration of 10µmol/L.[8,9] After 1hr, tubes were immediately centrifuged at 520 RCF room temperature for 15 minutes, before removal of supernatants, and resuspension of the cell pellets in Tri reagent (Cambridge Bioscience Ltd, Cambridge, UK). For all 6 donors, each lysed cell/Tri preparation was distributed to replicate tubes for RNA sequencing library generation, providing 96 paired samples.

#### RNA Sequencing analytics

RNA extraction, library preparations (48 for ribosomal (r)RNA depletion and paired 48 samples for polyA selection), alignments to GRCh38/hg38[10], unique gene read counts, differential expression (DESeq2)[11] and differential splice site (DEXSEq)[12] analyses were performed by Genewiz (Leipzig, Germany). Poly-A selected samples yielded 18,063,312-30,338,567 reads (median 22,354,247), 5,419-9,102 Mb sequence (median 6,707) with mean quality score 35.4-36.1 (median 36.0) and 91.9-95.0% (median 94.3%) bases exceeding a quality score of 35. All 48 GC distribution plots were acceptable, and passed quality control. There were also high-quality sequencing metrics for rRNA depleted samples: 5,918-24,259 (median 11,834) reads; 5,918-24,259 (median 11,834) Mb of sequence; mean quality scores 35.0-35.7 (median 35.5); 89.1-92.4% (median 91.3%) of bases exceeded quality score of 35, but 2 treatment libraries flagged after rRNA depletion also failed after a second round (eg GC content in *FigSA1)*.

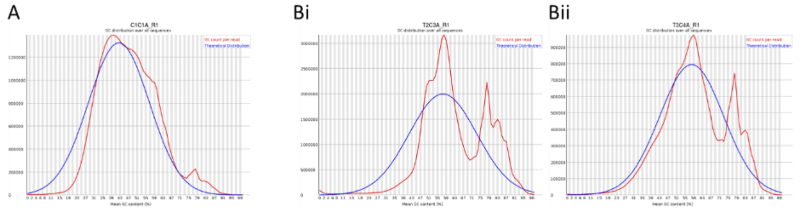

*FigSA1:* GC distribution in individual rRNA-depleted libraries (x axis: mean GC content, y axis number of sequences with content; blue curve: theoretical distribution, red curve: actual reads. **A)** Typical acceptable GC distribution observed in 46 of the 48 rRNA-depleted libraries (with higher matching seen in all 48 PolyA selected libraries). **B)** GC distributions in the two rRNA-depleted libraries failing quality control (and excluded from analysis) – 2 patients, 1 treatment.

Although the technical metrics for quality were met by the remaining 46 rRNA-depleted libraries, initial review of control, untreated samples indicated more marked *SMAD4* variability in rRNA-depleted samples than in poly A-selected samples (*Fig S3*). For rRNA depleted control untreated samples, the standard deviation represented 26.5% of the mean value (441/1662 normalised reads). We speculated that this may reflect different quantities of persistent rRNA.[13] To explore if variability could be reduced, normalisation was performed to the 25 genes with GINI coefficients (GC) <0.15 within multiple cell lines,[14,15] i.e. the least variable of all known human transcripts. First, within each library, the intra-assay coefficient of variation (CV)[16] was calculated for each replicate pair based on reads normalised by total reads per library, as *CV= 100 x standard deviation (SD)/mean)*. Most GINI genes met a 10% coefficient of correlation (CV10) between replicate rRNA-depleted libraries, but in three library pairs (two affected by samples in *FigSA1)*, fewer than 30% of GINI genes met CV10. The remaining libraries showed broadly consistent rankings of GINI genes (*Fig S4*). For the main text rRNA-depleted expression data presented for the 45 libraries meeting GINI-gene quality metrics, DeSeq2[11] normalisations were performed with the 25 GINI genes:[14,15] The ratio of alignment counts for each selected housekeeper gene in each monocyte dataset to the geometric mean of that gene across all datasets was calculated. Then, the median value of these ratios in each library was used to generate the 'size factor' to scale that library's alignments (*Table S8*). Following GINI normalisations, the standard deviation in the untreated samples from controls reduced to 5.0% of the mean (125/2488 reads).

#### Fig. S1. Design of comparison filter sets.

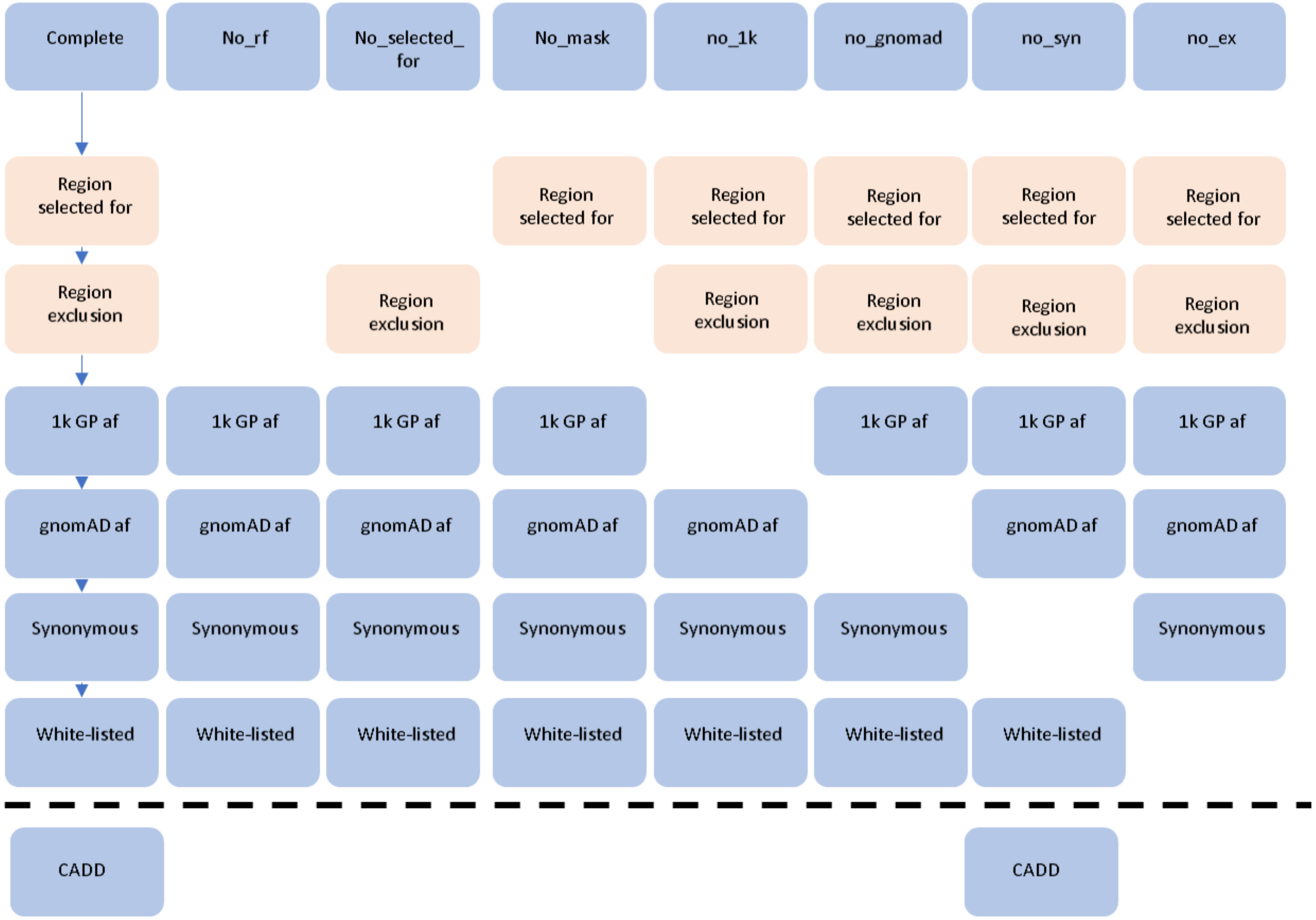

Design of comparison filter sets, with naming of the filter set based on the component it was missing**.** One complete filter set and 7 comparison filter sets lacking at least one component were designed. The Combined Annotation-Dependent Depletion (CADD) score[17] filter was not included in the comparison process as removal of some filters would leave too many variants to annotate CADD score. Under the dotted line, the CADD score filter was added to access the effect of the synonymous variant filter.

#### Fig. S2. Efficiency testing of GROFFFY in 98 whole genomes

**A**

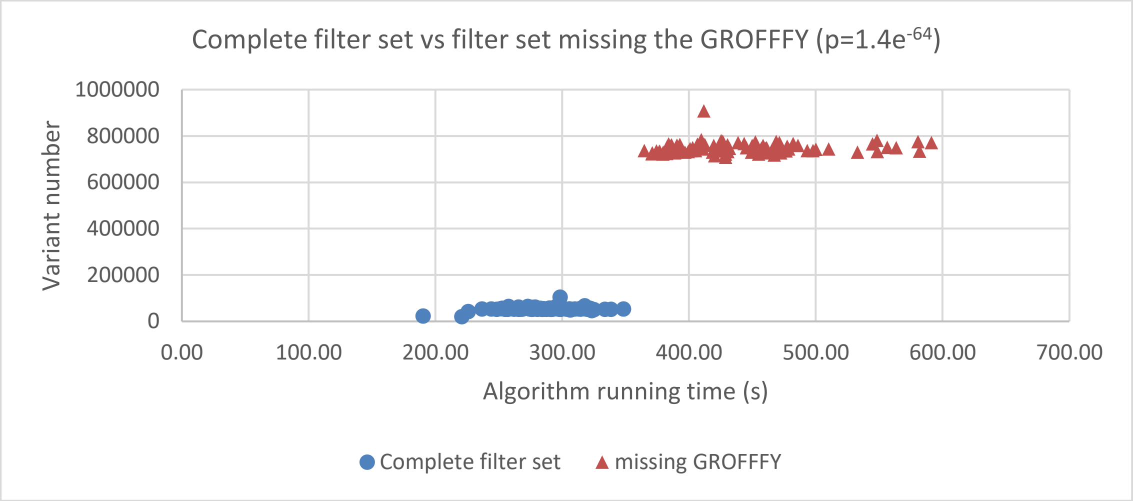

**B**

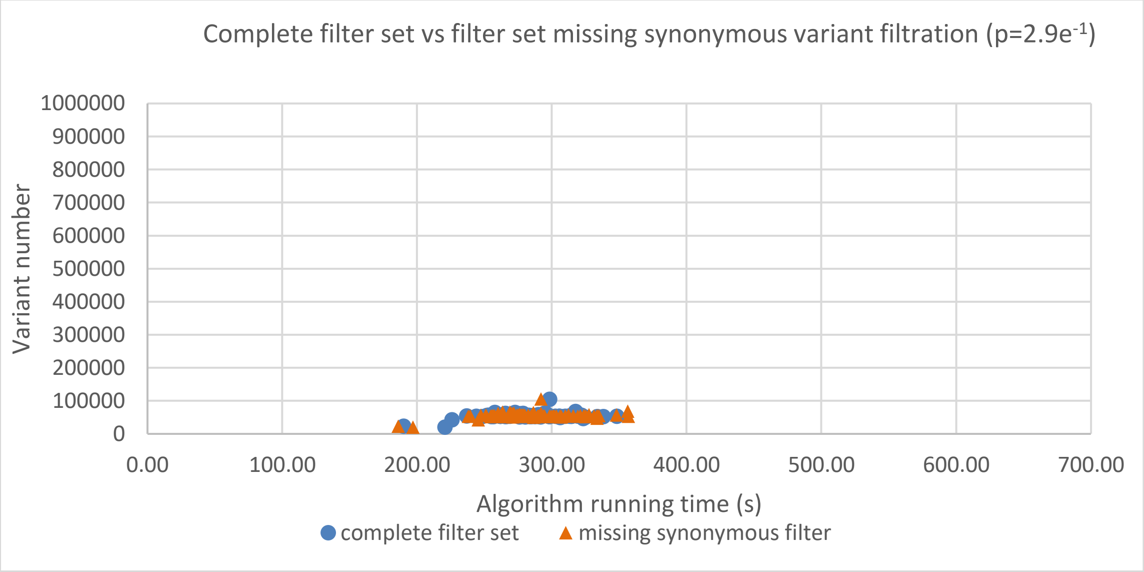

**A)** Efficiency of GROFFFY in reducing all dataset numbers and run times pre GROFFFY filter (red datasets) and post GROFFFY filter (blue datasets, $p=1.4e^{-64}$). **B)** Efficiency of synonymous filter in reducing dataset numbers and run times pre synonymous filter (orange datasets) and post synonymous filter (blue datasets, $p=2.9e^{-1}$). Contrast the lack of efficiency of the synonymous filter in **B),** to GROFFFY in **A)**.

#### Fig. S3: Differential *SMAD4* expression in control and 1hr stress conditions

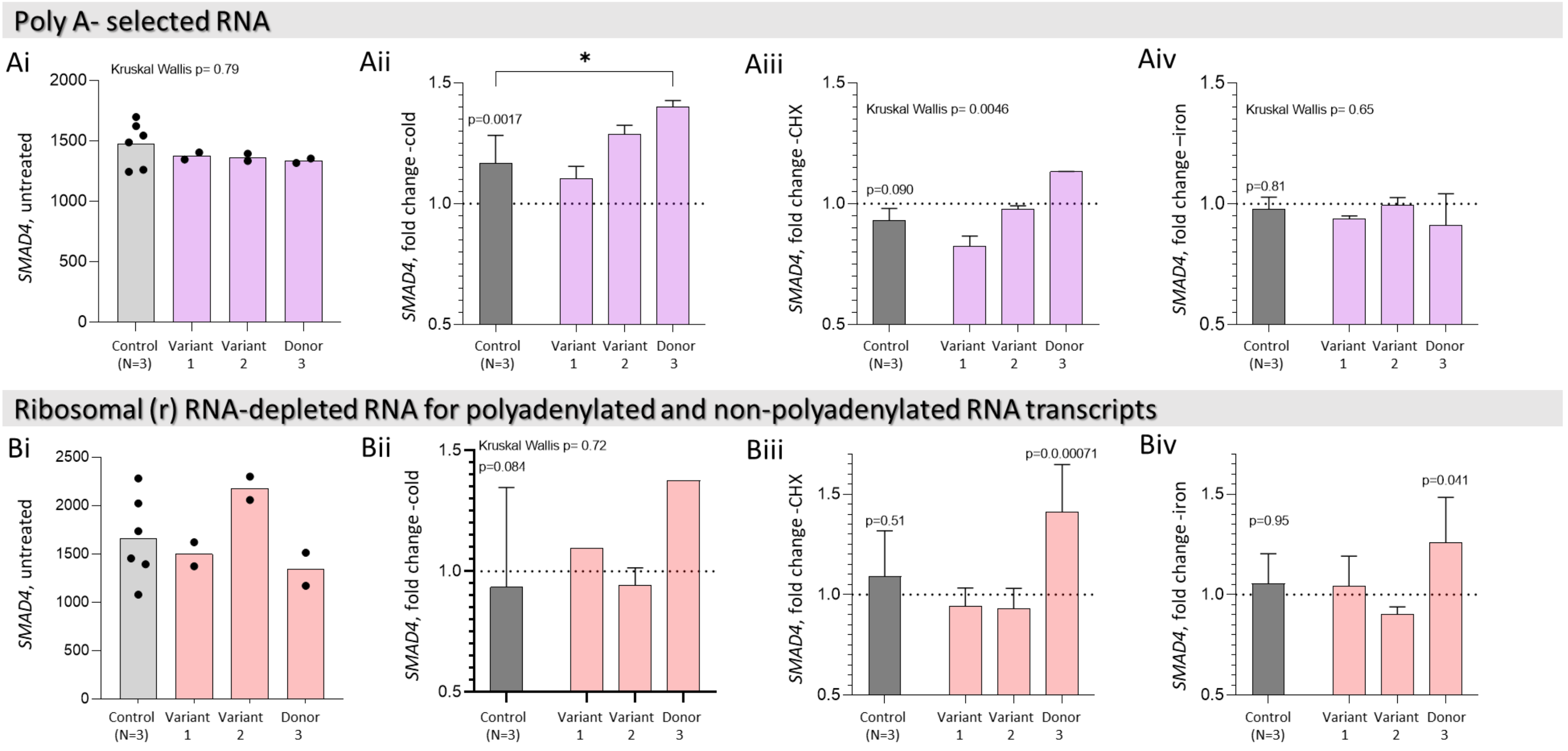

*SMAD4* expression in **A)** Poly A selected and **B)** ribosomal (r)RNA-depleted libraries, from 3 healthy controls and 3 patient donors under 4 conditions. **i)** Normalised *SMAD4* expression in untreated samples calculated by Genewiz. **iii-iv)** Fold change in normalised expression (treated/untreated per donor pair) after **ii)**1hr cold (32ºC); **iii)** 1hr cycloheximide (CHX); **iv)** 1hr ferric citrate (iron). **Ai)** In poly A-selected libraries, note similar intra-replicate values, with control standard deviation 12.7% of control mean (187/1476). **ii)** After a 1hr cold stress, control polyadenylated SMAD4 expression increased (Genewiz-calculated DeSeq2[11] Benjamini p=0.0017), with a further increase in Donor 3 (donor-based DeSeq2[11] Benjamini p=0.00059; pairwise p=0.036 calculated by Dunn's test post Kruskal Wallis for fold changes). There was no significant change in control polyadenylated *SMAD4* expression after **iii)** CHX or **iv)** ferric citrate. In **iii)**, the Kruskal Wallis p value for CHX (p=0.0046) suggested differences between the 4 groups but no pairwise p-values reached significance by Dunn's test. **Bi)** In rRNA-depleted libraries, there was greater variability between replicates and controls, with control untreated standard deviation 26.5% of mean (441/1662), limiting statistical comparisons. There was no change in control 'total' RNAs after stresses (**Bii-iv)**, but Donor 3 expression differences met Genewiz donor-based DESeq2[11] Benjamini significance after **iii)** CHX and **iv)** iron.

### Fig. S4. The 25 "GINI genes" in the rRNA-depleted libraries

A

| GINI gene | Number of pairs meeting CV10 | % of pairs meeting CV10 |
|---|---|---|
| *C2orf49* | 20 | 83.3 |
| *CHCHD4* | 9 | 37.5 |
| *CHTOP* | 22 | 91.7 |
| *CLINT1* | 20 | 83.3 |
| *CNOT2* | 16 | 66.7 |
| *CNOT4* | 14 | 58.3 |
| *COX4I1* | 17 | 70.8 |
| *HNRNPK* | 16 | 66.7 |
| *IK* | 20 | 83.3 |
| *INTS14* | 20 | 83.3 |
| *KAT5* | 18 | 75.0 |
| *NXF1* | 23 | 95.8 |
| *PARK7* | 22 | 91.7 |
| *PCBP1* | 21 | 87.5 |
| *PCBP2* | 19 | 79.2 |
| *RBM45* | 14 | 58.3 |
| *RPRD2* | 19 | 79.2 |
| *SF3B2* | 22 | 91.7 |
| *SNW1* | 18 | 75.0 |
| *SRP19* | 14 | 58.3 |
| *SUPT7L* | 21 | 87.5 |
| *TXNL1* | 18 | 75.0 |
| *UBE2Q1* | 23 | 95.8 |
| *UBR2* | 19 | 79.2 |
| *UXT* | 16 | 66.7 |

B

| GINI gene | Normalised read count | GINI gene colour coded |
|---|---|---|
| *HNRNPK* | 4272 | *HNRNPK* |
| *PCBP2* | 3997 | *PCBP2* |
| *UBR2* | 3708 | *UBR2* |
| *NXF1* | 2566 | *NXF1* |
| *CNOT2* | 2291 | *CNOT2* |
| *PCBP1* | 1586 | *PCBP1* |
| *SF3B2* | 1544 | *SF3B2* |
| *CHTOP* | 1490 | *CHTOP* |
| *CLINT1* | 1436 | *CLINT1* |
| *RPRD2* | 1410 | *RPRD2* |
| *COX4I1* | 1138 | *COX4I1* |
| *CNOT4* | 1079 | *CNOT4* |
| *IK* | 842 | *IK* |
| *SUPT7L* | 708 | *SUPT7L* |
| *UBE2Q1* | 695 | *UBE2Q1* |
| *SNW1* | 610 | *SNW1* |
| *PARK7* | 521 | *PARK7* |
| *TXNL1* | 488 | *TXNL1* |
| *C2orf49* | 486 | *C2orf49* |
| *INTS14* | 466 | *INTS14* |
| *KAT5* | 429 | *KAT5* |
| *UXT* | 281 | *UXT* |
| *SRP19* | 247 | *SRP19* |
| *RBM45* | 242 | *RBM45* |
| *CHCHD4* | 67 | *CHCHD4* |

C

| | | | | | | | | | | | | | |
|---|---|---|---|---|---|---|---|---|---|---|---|---|---|
| HNRNPK | ENSG00000165119 | 4271.959 | 4845.673 | 5132.413 | 4896.213 | 4893.896 | 5177.067 | 4477.034 | 4650.642 | 4331.849 | 4580.689 | 4562.294 | 4801.156 |
| PCBP2 | ENSG00000197111 | 3997.388 | 4185.219 | 4186.606 | 4046.257 | 3452.316 | 3556.738 | 4101.136 | 3969.546 | 3981.956 | 4233.9 | 3999.941 | 4457.828 |
| UBR2 | ENSG00000024048 | 3707.562 | 3902.168 | 3844.964 | 3757.74 | 3393.778 | 3376.607 | 3822.727 | 3814.843 | 3339.631 | 3182.559 | 3021.218 | 3086.328 |
| NXF1 | ENSG00000162231 | 2566.057 | 2483.39 | 2716.349 | 2697.245 | 2814.043 | 2864.253 | 2697.844 | 2694.751 | 2390.653 | 2540.56 | 2531.222 | 2584.471 |
| CNOT2 | ENSG00000111596 | 2290.638 | 2402.417 | 2205.086 | 2182.593 | 2299.193 | 2458.108 | 2291.949 | 2283.883 | 2167.631 | 2255.227 | 2022.185 | 2151.461 |
| PCBP1 | ENSG00000169564 | 1586.413 | 1758.158 | 1702.813 | 1748.258 | 2184.939 | 2294.971 | 1743.569 | 1694.202 | 1695.987 | 1696.633 | 1811.462 | 1808.133 |
| SF3B2 | ENSG00000087365 | 1544.041 | 1587.06 | 1678.239 | 1664.822 | 1924.693 | 1820.853 | 1594.522 | 1692.193 | 1658.437 | 1668.1 | 1726.41 | 1750.157 |
| CHTOP | ENSG00000160679 | 1489.804 | 1550.446 | 1669.248 | 1576.707 | 1746.964 | 1733.336 | 1474.534 | 1502.33 | 1496.86 | 1598.961 | 1652.784 | 1715.734 |
| CLINT1 | ENSG00000113282 | 1435.568 | 1532.14 | 1545.178 | 1501.069 | 1657.394 | 1696.8 | 1424.852 | 1457.627 | 1391.039 | 1477.146 | 1336.699 | 1390.523 |
| RPRD2 | ENSG00000163125 | 1410.145 | 1463.137 | 1350.982 | 1394.239 | 1517.044 | 1527.715 | 1360.171 | 1446.576 | 1341.542 | 1315.823 | 1246.57 | 1350.665 |
| COX4I1 | ENSG00000131143 | 1138.116 | 1193.463 | 1291.045 | 1164.985 | 1174.986 | 1192.943 | 1171.753 | 1233.608 | 1166.88 | 1189.618 | 1224.99 | 1323.488 |
| CNOT4 | ENSG00000080802 | 1078.795 | 1123.757 | 1114.23 | 1063.614 | 1079.774 | 1039.152 | 1059.265 | 1126.119 | 1080.402 | 1186.326 | 988.8778 | 1031.796 |
| IK | ENSG00000113141 | 842.3582 | 850.5628 | 912.8419 | 918.5761 | 951.4146 | 954.1845 | 831.476 | 947.3066 | 908.5845 | 924.0394 | 886.0548 | 985.5957 |
| SUPT7L | ENSG00000119760 | 707.6147 | 742.8342 | 758.2042 | 716.6141 | 935.1933 | 913.4001 | 747.1097 | 745.8909 | 724.8195 | 740.7679 | 726.1079 | 751.8791 |
| UBE2Q1 | ENSG00000160714 | 694.9031 | 717.4863 | 724.04 | 716.6141 | 830.1075 | 775.7528 | 679.6168 | 680.0918 | 699.2175 | 702.3578 | 672.7923 | 730.138 |
| SNW1 | ENSG00000100603 | 610.1588 | 691.4343 | 649.1187 | 604.3264 | 702.4529 | 713.7266 | 590.5635 | 644.9319 | 613.3088 | 665.045 | 582.6635 | 598.7856 |
| PARK7 | ENSG00000116288 | 521.1773 | 516.8155 | 549.0237 | 502.9555 | 593.8407 | 576.0793 | 525.8828 | 537.4433 | 508.0564 | 514.6965 | 515.3843 | 570.1006 |
| TXNL1 | ENSG00000091164 | 488.1271 | 497.8045 | 522.6514 | 499.8364 | 542.3557 | 550.5891 | 488.3867 | 536.941 | 487.0059 | 501.5273 | 502.6901 | 532.6565 |
| C2orf49 | ENSG00000135974 | 486.4322 | 471.7525 | 490.8848 | 484.2409 | 512.0289 | 543.7917 | 482.7623 | 488.7218 | 473.9204 | 492.7479 | 476.0323 | 502.7625 |
| INTS14 | ENSG00000138614 | 466.0935 | 451.3334 | 486.0898 | 466.306 | 464.7756 | 460.5236 | 472.4508 | 481.6898 | 465.3864 | 465.312 | 469.6852 | 479.2097 |
| KAT5 | ENSG00000172977 | 428.8061 | 414.0156 | 480.0961 | 426.5374 | 428.1013 | 454.5759 | 467.7638 | 451.5528 | 444.9049 | 460.9223 | 465.8769 | 461.0921 |
| UXT | ENSG00000126756 | 281.351 | 334.4514 | 294.2911 | 329.8452 | 302.5625 | 288.0397 | 284.9703 | 284.2924 | 324.2913 | 346.7891 | 336.3962 | 384.0924 |
| SRP19 | ENSG00000153037 | 247.4533 | 287.9803 | 245.7421 | 257.3261 | 222.8666 | 234.5102 | 281.2207 | 260.6851 | 255.4505 | 244.7278 | 232.3038 | 255.4577 |
| RBM45 | ENSG00000155636 | 242.3686 | 254.8872 | 230.1584 | 247.9688 | 217.9297 | 224.3141 | 228.7262 | 253.1509 | 250.3301 | 240.338 | 214.5319 | 215.5991 |
| CHCHD4 | ENSG00000163528 | 66.94798 | 80.97245 | 50.34716 | 68.62028 | 87.45398 | 72.22233 | 72.17999 | 73.83567 | 63.72039 | 59.26144 | 62.20155 | 49.82331 |

| | | | | | | | | | | | | |
|---|---|---|---|---|---|---|---|---|---|---|---|---|
| HNRNPK | 4315.589 | 5017.544 | 4189.636 | 4114.252 | 4670.316 | 4515.61 | 4524.017 | 4299.934 | 4806.555 | 3934.802 | 4570.657 | 4686.674 |
| PCBP2 | 3403.313 | 3656.47 | 3656.516 | 4028.27 | 4386.249 | 4345.601 | 4482.139 | 4205.628 | 4106.635 | 3497.184 | 3417.977 | 3472.409 |
| UBR2 | 2880.87 | 2747.073 | 3089.268 | 2780.094 | 3572.227 | 3429.21 | 3692.44 | 3482.61 | 3584.934 | 3090.914 | 2863.958 | 2934.56 |
| NXF1 | 2454.455 | 2633.776 | 2655.005 | 2659.719 | 2558.136 | 2355.249 | 2710.103 | 2750.611 | 2625.783 | 2152.981 | 2862.813 | 2856.551 |
| CNOT2 | 2250.965 | 2525.011 | 2304.3 | 2310.057 | 2200.76 | 2066.026 | 2425.333 | 2213.961 | 2074.92 | 2066.461 | 2448.443 | 2528.094 |
| PCBP1 | 2105.779 | 2350.534 | 1950.064 | 1974.727 | 2075.526 | 1916.75 | 1760.071 | 1854.698 | 1843.773 | 1744.203 | 2401.512 | 2384.394 |
| SF3B2 | 1706.801 | 1776.496 | 1628.78 | 1807.061 | 1855.603 | 1915.713 | 1634.437 | 1609.949 | 1544.578 | 1721.633 | 1817.732 | 1744.929 |
| CHTOP | 1669.075 | 1646.582 | 1554.637 | 1546.248 | 1715.096 | 1901.2 | 1573.415 | 1494.311 | 1531.617 | 1623.827 | 1656.334 | 1653.577 |
| CLINT1 | 1624.491 | 1625.433 | 1315.734 | 1342.757 | 1472.264 | 1455.444 | 1488.463 | 1432.563 | 1450.608 | 1324.14 | 1605.968 | 1558.119 |
| RPRD2 | 1306.68 | 1392.042 | 1206.286 | 1289.734 | 1426.447 | 1394.283 | 1365.222 | 1159.747 | 1313.432 | 1281.507 | 1500.659 | 1551.96 |
| COX4I1 | 991.1564 | 1173.756 | 1116.844 | 1119.203 | 1081.29 | 1054.264 | 1093.613 | 1153.011 | 1139.532 | 1225.08 | 1222.505 | 1292.274 |
| CNOT4 | 990.0132 | 993.9918 | 996.8038 | 965.8677 | 977.437 | 934.0139 | 1090.024 | 1048.6 | 1093.086 | 827.5875 | 995.8606 | 973.0545 |
| IK | 834.5377 | 919.9712 | 819.0973 | 816.8317 | 855.2574 | 893.5849 | 778.9303 | 751.0852 | 792.8116 | 761.1297 | 881.3939 | 853.9888 |
| SUPT7L | 826.5353 | 917.7052 | 693.1729 | 690.7244 | 836.9304 | 860.4123 | 684.4057 | 647.7969 | 704.2414 | 635.7377 | 869.9472 | 814.9845 |
| UBE2Q1 | 690.4942 | 712.2601 | 622.5611 | 663.4967 | 656.7155 | 664.4871 | 668.851 | 641.0607 | 627.5525 | 628.2141 | 685.6557 | 707.2095 |
| SNW1 | 683.635 | 697.1539 | 611.9693 | 630.5368 | 626.1706 | 642.7176 | 656.8859 | 635.4472 | 592.9885 | 574.2956 | 669.6304 | 669.2316 |
| PARK7 | 579.6036 | 574.0378 | 583.7245 | 556.0188 | 583.4077 | 601.252 | 570.7369 | 561.3491 | 582.1873 | 536.6779 | 620.4097 | 580.9587 |
| TXNL1 | 505.2954 | 542.3147 | 556.6567 | 530.2241 | 531.4814 | 529.7237 | 504.9287 | 479.3922 | 495.7773 | 478.9976 | 562.0317 | 556.3244 |
| C2orf49 | 480.145 | 495.4853 | 429.5554 | 428.4784 | 484.1368 | 443.6825 | 477.4089 | 452.4474 | 477.4152 | 473.9819 | 481.905 | 533.743 |
| INTS14 | 474.429 | 484.1556 | 422.4942 | 418.4471 | 445.9556 | 395.997 | 417.5832 | 435.6069 | 465.5338 | 466.4584 | 456.7223 | 484.4744 |
| KAT5 | 377.2568 | 399.5606 | 398.9569 | 371.1569 | 424.5742 | 383.5573 | 362.5436 | 396.3125 | 451.4922 | 452.6653 | 424.6716 | 448.5494 |
| UXT | 289.2302 | 339.8908 | 278.9168 | 272.2772 | 262.6862 | 258.1237 | 275.1981 | 273.9384 | 330.5182 | 313.4801 | 309.0602 | 324.3515 |
| SRP19 | 252.6477 | 271.9127 | 196.5363 | 236.4513 | 167.997 | 203.1817 | 248.8748 | 220.0489 | 252.7492 | 231.9753 | 247.2481 | 260.7129 |
| RBM45 | 186.342 | 265.1148 | 124.7476 | 151.902 | 146.6156 | 125.4336 | 204.6038 | 190.8587 | 250.589 | 211.9126 | 233.5121 | 229.9201 |
| CHCHD4 | 72.02175 | 68.73348 | 58.8432 | 37.25899 | 70.25328 | 81.89467 | 55.03962 | 72.97539 | 45.36524 | 51.41074 | 75.54804 | 59.53287 |

| | | | | | | | | | | |
|---|---|---|---|---|---|---|---|---|---|---|
| HNRNPK | 4544.765 | 4294.074 | 4717.518 | 4887.922 | 4406.852 | 4283.877 | 4794.149 | 4455.053 | 4099.369 | 4367.944 |
| PCBP2 | 4089.91 | 4011.856 | 3797.782 | 3651.086 | 4184.448 | 4224.325 | 4013.368 | 3981.501 | 4012.369 | 4187.711 |
| UBR2 | 3491.318 | 3405.676 | 3254.357 | 3325.96 | 4081.383 | 4048.552 | 3481.546 | 2913.19 | 3966.797 | 4015.357 |
| NXF1 | 2513.522 | 2433.793 | 3156.566 | 3135.808 | 2692.714 | 2694.232 | 2503.148 | 2424.135 | 2657.651 | 2703.496 |
| CNOT2 | 2315.882 | 2216.004 | 2855.763 | 2741.922 | 2565.781 | 2573.208 | 2279.376 | 2018.233 | 2304.471 | 2440.533 |
| PCBP1 | 1720.126 | 1807.649 | 2130.372 | 2146.849 | 1605.648 | 1641.513 | 1819.239 | 1765.954 | 1644.719 | 1944.154 |
| SF3B2 | 1602.867 | 1620.714 | 1615.419 | 1767.394 | 1596.968 | 1494.554 | 1718.493 | 1678.573 | 1418.933 | 1513.761 |
| CHTOP | 1561.258 | 1510.912 | 1611.705 | 1718.158 | 1557.912 | 1483.989 | 1468.565 | 1541.863 | 1376.468 | 1436.94 |
| CLINT1 | 1518.704 | 1495.485 | 1489.156 | 1528.855 | 1493.903 | 1456.134 | 1426.911 | 1375.556 | 1281.182 | 1337.467 |
| RPRD2 | 1507.357 | 1469.169 | 1464.399 | 1387.09 | 1434.234 | 1328.386 | 1314.54 | 1152.874 | 1141.361 | 1239.964 |
| COX4I1 | 1217.99 | 1084.408 | 1166.072 | 1171.471 | 1149.991 | 1132.442 | 1024.896 | 1096.498 | 1025.36 | 1083.368 |
| CNOT4 | 1091.273 | 1019.979 | 892.5034 | 951.6082 | 885.276 | 938.4188 | 915.4315 | 841.4003 | 900.0384 | 1029.2 |
| IK | 838.7863 | 817.6164 | 835.5615 | 865.0212 | 815.8426 | 897.1168 | 853.4341 | 790.6626 | 654.5734 | 759.3427 |
| SUPT7L | 691.2658 | 634.3106 | 824.4207 | 852.2878 | 697.5888 | 706.9358 | 688.7533 | 651.1339 | 618.3233 | 679.5674 |
| UBE2Q1 | 682.755 | 625.2361 | 688.2551 | 713.0695 | 669.3815 | 695.4096 | 653.8797 | 615.8994 | 612.109 | 614.5653 |
| SNW1 | 668.5703 | 617.9765 | 658.5462 | 686.7538 | 662.8721 | 626.2529 | 622.8809 | 601.8056 | 548.9302 | 576.1549 |
| PARK7 | 600.484 | 557.177 | 561.9925 | 564.5134 | 535.9391 | 520.5967 | 552.1651 | 541.2022 | 540.6445 | 543.6539 |
| TXNL1 | 486.061 | 508.1745 | 559.5167 | 555.1755 | 493.6281 | 509.0706 | 527.9473 | 536.9741 | 483.68 | 477.6669 |
| C2orf49 | 476.6046 | 470.0614 | 554.5652 | 521.2199 | 491.4584 | 508.1101 | 493.0737 | 460.8675 | 447.4299 | 440.2415 |
| INTS14 | 473.7677 | 457.357 | 485.2446 | 499.9975 | 451.3172 | 445.6769 | 356.4855 | 357.9827 | 363.5368 | 383.1184 |
| KAT5 | 460.5286 | 454.6347 | 415.9239 | 455.0063 | 430.7041 | 444.7164 | 319.6745 | 294.5606 | 346.9653 | 346.6778 |
| UXT | 365.9642 | 330.3134 | 262.4282 | 323.428 | 309.1957 | 307.3634 | 235.3967 | 181.8101 | 226.8221 | 235.3864 |
| SRP19 | 248.7044 | 238.6605 | 243.8602 | 251.2721 | 291.8373 | 282.3901 | 176.3053 | 167.7163 | 194.7149 | 185.1575 |
| RBM45 | 244.9218 | 231.4009 | 174.5395 | 190.1519 | 263.63 | 216.1149 | 152.0876 | 132.4818 | 169.8576 | 177.2784 |
| CHCHD4 | 53.90171 | 56.26217 | 50.75262 | 53.48021 | 54.24485 | 61.47267 | 49.40424 | 52.14709 | 52.82158 | 70.91138 |

The 25 genes with lowest GINI coefficients of variation ("GINI genes")[14,15] in the rRNA-depleted libraries.

**A)** Number and percentage (%) of replicates meeting CV10 for each gene.

**B)** Relative expression (normalized to total library read counts) for GINI genes in first rRNA control untreated library. The third column indicates colour code used in C).

**C)** Relative expression of the GINI genes across representative rRNA-depleted libraries meeting GINI gene quality control. Note the general consistency in relative expression across the 24 donor-treatment combinations.

#### Fig. S5. *SMAD4* last exon transcripts, regions, and differential use in volunteer and patient monocytes after 1hr cycloheximide

A

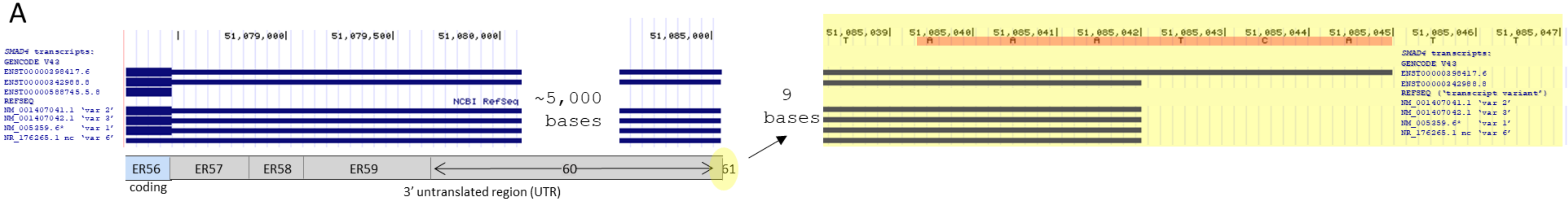

B

| | Exon | Region | Start | End | Bases |
|---|---|---|---|---|---|
| 22- SMAD4 Final exon | | ER56 | 51,078,256 | 51,078,467 | 212 |
| | | ER57 | 51,078,468 | 51,078,870 | 403 |
| | | ER58 | 51,078,871 | 51,079,143 | 273 |
| | | ER59 | 51,079,144 | 51,079,777 | 634 |
| | | ER60 | 51,079,778 | 51,085,039 | 5262 |
| | | ER61 | 51,085,040 | 51,085,045 | 6* |

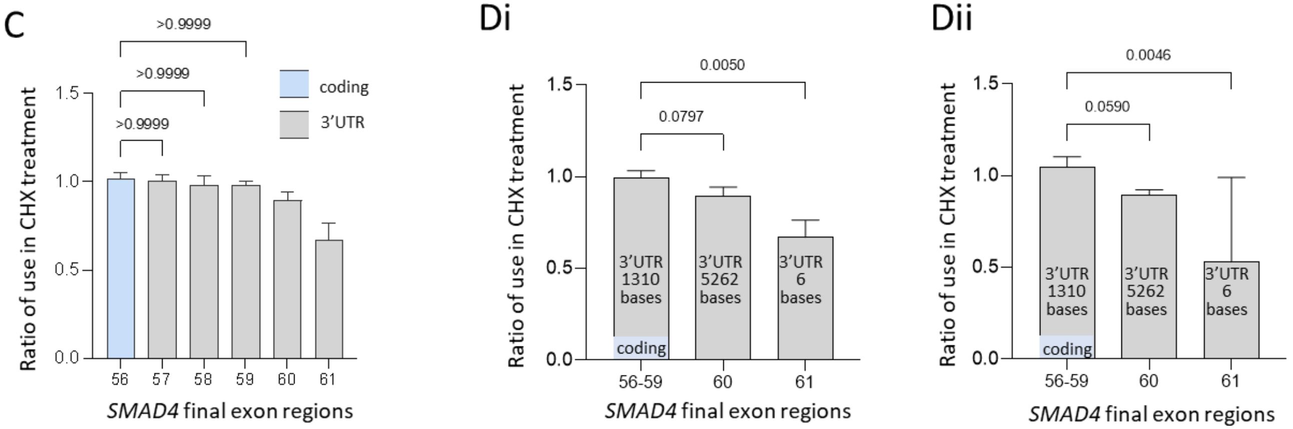

**A)** Annotated screen shot of last *SMAD4* exon (GRChg38/hg38[10] chr18:51,078252-51,085,050), as denoted on the University of California Santa Cruz (UCSC) Genome Browser[18] by kilobase (left) and individual base (right, last 9 nucleotides highlighted). REFSEQ[19] transcripts (as in main manuscript Fig.3) are now shown with three additional GENCODE[20] transcripts: All distinguish coding regions (thick bars) from untranslated regions (thin bars). Lower bar indicates DEXSeq[12] *SMAD4* exon regions (ERs). ER56 is denoted in pale blue for coding sequence, though while present in all 7 transcripts shown, it encodes inframe coding sequence only in 6 of the transcripts. ER57-61 are untranslated regions (grey) contained in 6 of the 7 transcripts but ER61 comprises 6 nucleotides highlighted in red, and is present in full only in GENCODE transcript ENST00000398417.6. **B)** DEXSeq[12] division of *SMAD4* last exon into the 6 contiguous but separately-examined regions between chr18:51,078,256-51,085,045. The natural stop codon at chr18:51,078,465 separates the final coding nucleotides (ER56, blue) from the 3' UTR (ER57-ER61). The final AAUAAA site is at chr18:51,083,978 in the penultimate region ER60. **C)** Differential use of these last exon regions after 1hr CHX 100µg/mL treatment in the control donors, calculated as DEXSeq[12] reads after CHX, to reads without CHX (3 donors, 2 replicates per donor). Mean and standard deviation displayed, overall Kruskal Wallis p=0.0001, displayed pairwise p values calculated by Dunn's test. Note CHX treatment did not modify use of the first 4 regions spanning 212 coding and 1,310 noncoding bases, with all ratios ~1.0. **D)** Comparison of treatment/untreated ratios for 1hr CHX 100µg/mL (calculated as in **C**) for penultimate (ER60) and final (ER61) exon regions, compared to the now pooled ER56-59 providing proximal 1,522 bases in **i)** the 3 healthy donors and **ii)** the 3 patients. Note in both volunteers and patients, there was less use of ER61 (Dunn's p≤0.005), and a trend to lower use of ER60 (Dunn's p≤0.08), after 1hr CHX suggesting shorter 3'UTRs, as would be predicted,[1] and potentially, specifical reduction of transcripts such as ENST00000398417.6 that encode the final ER61 region.

#### Fig. S6. Individual *SMAD4* exon region use in monocytes from 6 volunteers and patients

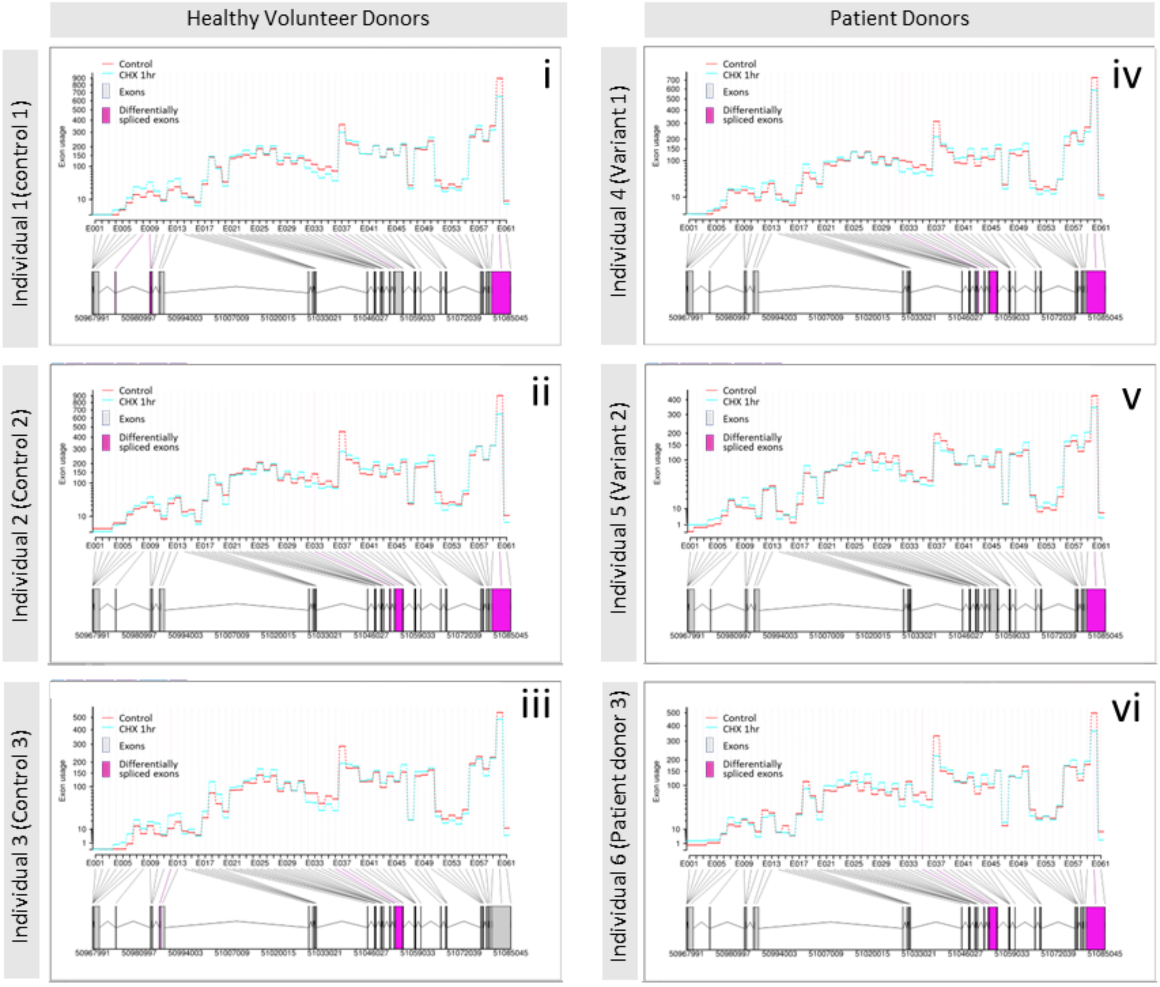

**Differential use of *SMAD4* exon regions in monocytes after 1hr cycloheximide (CHX).** Individual DEXSeq[12] images of *SMAD4* exon region use in monocytes with and without 1hr treatment with cycloheximide 100µg/mL (in the other two treatments of hypothermia and iron, *SMAD4* did not meet DEXSeq thresholds for differential exon use). Data are for 61 exon regions (ERs) in 22 exons, from **i-iii)** the 3 healthy volunteers; **iv-vi)** the 3 patients. In each case: *Upper graphs*: reads for exon regions untreated ('control', red lines) and 1hr CHX-treated (blue lines). *Lower graphs*: Genomic origin of exons and regions. DEXSeq[12] graphics denote exon regions that meet differential expression significance in purple. Note that although the trends to lower use of the penultimate, AAUAAA-containing region (ER60) did not meet statistical significance in in-house aggregated data compared only to the immediately preceding exon (*Fig S5*), for individual donors across all exon regions, DEXSeq[12] metrics denoted differential use in 5 of the 6 donors between resting and CHX-treated states. In each case there was lower use of ER60 after 1hr CHX suggesting shorter 3'UTRs, as would be predicted.[1]

#### Fig S7: *SMAD4* DNA variants in previously unsolved "SMAD4 positive control"

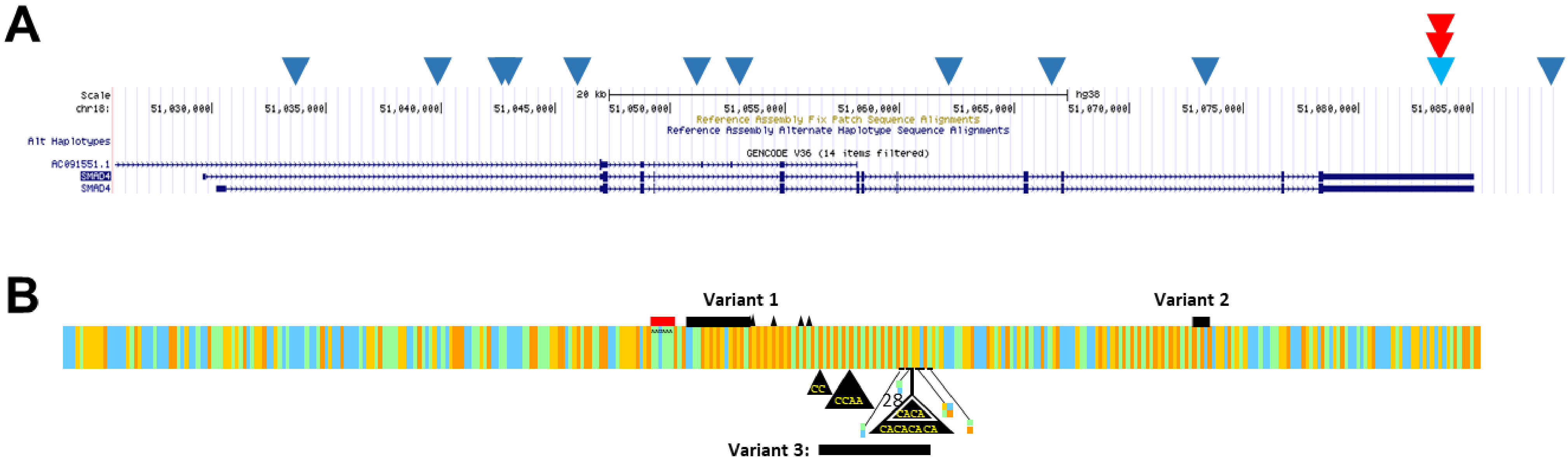

The previously unsolved clinical *SMAD4* case was noted to have similar expression patterns of total *SMAD4* expression, and ER60 region expression, to the Variant 1 donor (compare Fig. 7Ai/iii, Fig. 7Ci/iii and *Fig. S6iv/vi*). Blinded to the findings and project, a new team member was asked to examine their 100,000 Genomes Project bam file and check the "*number of coding, noncoding transcribed (eg UTRs), and intronic variants"* as indicated in *Table S7*. Multiple intronic DNA variants and a single exonic variant were identified. The exonic variant was reviewed and identified to be in the 3'UTR and sited adjacent to Variant 1. **A)** Variant sites shown superimposed on screenshots of GRCh38 from the University of California Santa Cruz (UCSC) Genome Browser. Dark blue inverted triangles indicate the sites of the patient's intronic and intergenic DNA variants, and bright blue, the sole exonic variant ("Variant 3") sited between the GROFFFY-solved Variants 1 and 2 (indicated by red inverted triangles). **B)** Cartoon of whole genome sequencing data indicating site of Variant 3, disrupting the (CA)n repeat element and challenging to sequence**.** Black triangles indicate sites of inserted bases; colour bases indicate single nucleotide substitutions, linked to their DNA sites demarcated by black horizontal bars. We speculate that the complexity of the reads in a repetitive region is the reason why the variant was not captured by the variant call format (vcf) file used as the basis for GROFFFY: Of 35 reads at the Variant 3, only 21 (60%) were complete with 14 $(CA)_{18}$ reads, 7 $(CA)_{20}$ reads. In comparison, of 43 reads at chr18:51,088,009 downstream of the *SMAD4* 3'UTR, 37 (86%) were complete and split 21:16 across the heterozygous alleles.

#### SUPPLEMENTARY TABLES

##### Table S1. Genomic region data used to create the GROFFFY.

| Name | Source | Format | Version |
|---|---|---|---|
| DNA-binding ChIP-seq | https://www.encodeproject.org/search/?type=Experiment&status=released | .bed | NA |
| DNase-seq | https://www.encodeproject.org/search/?type=Experiment&status=released | .bed | NA |
| CpG islands | http://hgdownload.cse.ucsc.edu/goldenpath/hg38/database | .txt | NA |
| Human genome annotation | ftp://ftp.ebi.ac.uk/pub/databases/gencode/Gencode_human/release_31/ | .gff3 | V31 |
| lncRNA (GENCODE) | https://www.gencodegenes.org/human/ | .gff3 | V31 |
| lncRNA (Lncipedia) | https://lncipedia.org/download | .bed | V5.2 |
| miRNA | http://www.mirbase.org/ftp.shtml | .gff3 | release 22.1 |
| ENCODE blacklist | https://www.encodeproject.org/annotations/ENCSR636HFF/ | .bed | NA |
| RepeatMasker | http://hgdownload.cse.ucsc.edu/goldenpath/hg38/database/ | .txt | NA |

**Genomic region data used to create the GROFFFY.** Names, download link, downloaded format and data version for the data used to create the GROFFFY are listed.

##### Table S2. List of options used in variant annotation.

| Option | Description |
|---|---|
| --vcf | Output in vcf format. Annotated information will be stored in INFO filed. Different annotated categories are separated by '\|'. |
| --pick | Pick one consequence annotation per variant. Consequences are chosen based on canonical status, APPRIS isoforms, biotype of the transcript, deleteriousness, transcript support level and etc. |
| --offline | Enable offline mode |
| --cache | Enable use of cache. Cache directory and version (V93) was also specified. |
| --assembly GRCh38 | Use genome assembly GRCh38 |
| --variant_class/af/af_gnomad | Annotation flags (sequence ontology variant class/global allele frequency in 1000 genome project/allele frequency from gnomAD database). |
| --force | Force overwrite if file already exits |

**A list of options used in variant annotation.** Options used to allow VEP to run in an offline mode and their descriptions were listed. Annotation flags were also added to optimize the program runtime by only including the interested information.

#### Table S3. ENCODE DNA binding call sites

- see end of document

#### Table S4. ENCODE DNase hypersensitivity sites (rDHSs)

- see end of document

#### Table S5. Variant numbers

| A) Filter applied sequentially | Number of Variants | Standard error of the mean (SEM) |
|---|---|---|
| Original | 4,867,167.11 | 4,961.82 |
| GROFFFY filter | 2,055,152.15 | 19,011.58 |
| Allele frequency (1000 GP) | 335,426.81 | 3,414.52 |
| Allele frequency (gnomAD) | 328,638.47 | 3,345.75 |
| Synonymous variants | 328,085.12 | 3,339.73 |
| Variants from the white list | 55,756.20 | 628.89 |
| CADD PHRED (<10) | 21,864.20 | 181.95 |

| B) Filter comparison | Number of Variants | Standard error of the mean (SEM) |
|---|---|---|
| Complete | 54842.78 | 786.28 |
| No_GROFFFY | 747042.62 | 2381.19 |
| No_positive | 65886.01 | 636.05 |
| No_mask | 676736.27 | 2166.04 |
| No_1000 genomes frequency | 1760169.16 | 1726.23 |
| No_gnomAD frequency | 61414.8 | 637.29 |
| No_synon | 56318.99 | 629.37 |
| No_white list | 333363.95 | 817.6 |

**A)** Variant numbers after applying each filter. Mean variant numbers for 98 patients were calculated after each filter was applied to the variant calling files. **B)** Variant numbers after applying comparison filter sets. Mean variant numbers for 98 patients after 7 comparison filter sets and 1 complete filter set applied to genomes were recorded.

#### Table S6. Wildtype and variant sequences for iFoldRNA

| cDNA sequence | Descriptor | cDNA Sequence* |
|---|---|---|
| NM_005359:7561_7920 | WT full region as seen in Figure 4a | ORIGIN NM_005359<br>7561 tttgaggggg ctttttactt atttccatgt tattcaaagg agactaggct tgatatttta<br>7621 ttactgttct tttatggaca aaaggttaca tagtatgccc ttaagactta attttaacca<br>7681 aaggcctagc accaccttag gggctgcaat aaacacttaa cgcgcgtgcg cacgcgcgcg<br>7741 cgcacacaca cacacacaca cacacacaca cacaggtcag agtttaaggc tttcgagtca<br>7801 tgacattcta gcttttgaat tgcgtgcaca cacacacgca cgcacacact ctggtcagag<br>7861 tttattaagg ctttcgagtc atgacattat agcttttgag ttggtgtgtg tgacaccacc |
| **gDNA Variant 1:** | **Descriptor** | **RNA Sequence*** |
| 18:51083986:CTTAACGCGCGTGCGCA>C<br>18:51084003:C>A<br>18:51084004:G>A<br>18:51084011:C>T<br>18:51084016:A>G<br>18:51084018:A>G | WT | ACUUAAUUUUAACCAAAGGCCUAGCACCACCUUAGGGGCUGCAAUAAACACUUAACGCGCGUGCGCACGCGCGCGCG<br>CACACACACACACACACACACACACACACACACAGGUCAGAGUUUAAGGCUUUCGAGUCAUGACAUUCUAGCUUU |
| 18:51083986:CTTAACGCGCGTGCGCA>C<br>18:51084003:C>A<br>18:51084004:G>A<br>18:51084011:C>T<br>18:51084016:A>G<br>18:51084018:A>G | Variant | ACUUAAUUUUAACCAAAGGCCUAGCACCACCUUAGGGGCUGCAAUAAACACAACGCGCGUGCACGCGCACACACACAC<br>ACACACACACACACAGGUCAGAGUUUAAGGCUUUCGAGUCAUGACAUUCUAGCUUU |
| **gDNA Variant 2:** | **Descriptor** | **RNA Sequence*** |
| 18:51084116:ACACT>A | WT | UUUAAGGCUUUCGAGUCAUGACAUUCUAGCUUUUGAAUUGCGUGCACACACACACGCACGCACACACUCUGGUCAG<br>AGUUUAUUAAGGCUUUCGAGUCAUGACAUUAUAGCUUUUGAGUUGGUGUGUGUGACACCACCCUCCUAAGUGGUG<br>UGUGCUUGUAAUUUUUUUUUU |
| 18:51084116 :ACACT>A | Variant | UUUAAGGCUUUCGAGUCAUGACAUUCUAGCUUUUGAAUUGCGUGCACACACACACGCACGCACACUGGUCAGAGUU<br>UAUUAAGGCUUUCGAGUCAUGACAUUAUAGCUUUUGAGUUGGUGUGUGUGACACCACCCUCCUAAGUGGUGUGUG<br>CUUGUAAUUUUUUUUUU |

Wildtype cDNA sequences (FASTA format), and wildtype/variant sequences as aligned to GRCh 38 genome build.[10] Wildtype sequences of nucleotides deleted in a variant are highlighted in green, with nucleotides that are substituted underlined. Variant sequences are in red, with the AAUAAA hexamer highlighted in yellow, and deletion-adjacent nucleotides in blue for reference. RNA sequences were those modelled in iFoldRNAv2,[21-23] centred on the variant. *Note uridine replaces thymidine in RNA.

#### Table S7. 100,000 Genomes Project Airlock transfers and Clinical Collaboration Requests

| A) AIRLOCK TRANSFERS (Project RR42: HHT-Gene-Stop) | | |
|---|---|---|
| **File name** | **Approval Date** | **Notes** |
| sizes 2.csv/sizes.csv | 2019/08/23 and 2019/08/27 | Number of variants after each filter |
| al_log.txt | 2019/10/07 | Number of variants after applying the comparison filter set (each removing one component), also includes synonymous filter comparison |
| time.py/vcf_af_filter.py | 2019/10/09 | Scripts for establishing the filter set and comparison |
| table.png/table_log2.png/table_log.png | 2019/10/11 and 2019/11/01 | High resolution graph for number of variants after each filter, also in log scale |
| validation.tsv | 2019/10/23 | Filtered variants in selected 5 genes of 34 patients |
| AIRLOCK_TRANSFER_(NAME)_2 | 2023/04/27 | Details of SMAD4 variants for previously unsolved case: Followed 2023/04/12 senior author request for Research Environment examination of *SMAD4* in specific GEL ID: *"….. when you are in the Research Environment, could you please look for SMAD4 variants in [GEL ID]? In the first instance, this would just be the number of coding, noncoding transcribed (eg UTRs), and intronic variants – it might be 0 – 0 – 0 or 150"* |
| **B) CLINICIAN COLLABORATION REQUESTS** | | |
| **Variant site** | **Date** | **Notes** |
| Chr18:51083986C (Variant 1) | 2022/09/12 | Enabled contact for resampling |
| Chr18: 51084116 A (Variant 2) | 2022/09/12 | Enabled contact for resampling |
| Chr18: 51084012G | 2023/04/24 | Details of single SMAD4 3'UTR variant as also in airlock transfer 2023/04/27. |

#### Table S8. RNA Sequencing Library Size Factors

| rRNA Depleted Library | Size factor |
|---|---|
| C1C1A | 1.138075 |
| C1C2A | 1.44176 |
| C1C3A | 1.681162 |
| C1C4A | 1.319289 |
| C1C5A | 1.4643 |
| C1C6A | 1.23388 |
| C1C7A | 1.029778 |
| C1C8A | 1.99758 |
| C2C1A | 1.690271 |
| C2C2A | 0.904089 |
| C2C3A | 0.762185 |
| C2C4A | 1.134408 |
| . | . |
| C2C6A | 1.387672 |
| C2C7A | 0.967989 |
| C2C8A | 0.784336 |
| C3C1A | 1.541809 |
| C3C2A | 0.677342 |
| C3C3A | 0.777739 |
| C3C4A | 0.631416 |
| C3C5A | 0.626968 |
| C3C6A | 0.901622 |
| C3C7A | 0.810988 |
| C3C8A | 0.829579 |
| T1C1A | 0.475731 |
| T1C2A | 1.008012 |
| T1C3A | 0.931979 |
| T1C4A | 0.704232 |
| T1C5A | 0.881458 |
| T1C6A | 0.994901 |
| T1C7A | 1.045663 |
| T1C8A | 1.062598 |
| T2C1A | 0.852478 |
| T2C2A | 0.913042 |
| . | . |
| T2C4A | 0.716733 |
| T2C5A | 0.793025 |
| T2C6A | 1.200489 |
| T2C7A | 0.899117 |
| T2C8A | 1.005649 |
| T3C1A | 1.123842 |
| T3C2A | 0.554218 |
| T3C3A | 0.815862 |
| . | . |
| T3C5A | 0.945863 |
| T3C6A | 0.575727 |
| T3C7A | 0.829579 |
| T3C8A | 0.91337 |

'Size factor' to scale individual library's alignments for DeSeq2[11] normalisations calculated using the 25 GINI genes for rRNA depleted libraries (N=45, note 3 libraries failed QC and are represented by ".") In each case, the ratio of alignment counts for each selected housekeeper gene in each monocyte dataset to the geometric mean of that gene across all datasets was calculated, then, the median value of these ratios in each library was used to generate the size factor indicated.
